# Long-Read Haplotype Phasing Resolves Allelic Configuration as a Missing Layer of Precision Oncology

**DOI:** 10.64898/2026.05.05.26351600

**Authors:** Josh N. Vo, Yi-Mi Wu, Rui Wang, Tiffany Pham, Xuhong Cao, Sophie Yeung, Mingyu Park, Yelena Kleyman-Smith, Guo Ci Teo, Alyssa J. Wu, Anne Li, Jamie C. Estill, Lakshmi P. Kunju, Chen Yang, Dan R. Robinson, Arul M. Chinnaiyan

**Author notes:** Co-corresponding authors: Dan R. Robinson, Michigan Center for Translational Pathology, University of Michigan Medical School, 1500 E. Medical Center Dr., Rogel Cancer Center, Ann Arbor, MI 48109-0940., Arul M. Chinnaiyan, Michigan Center for Translational Pathology, University of Michigan Medical School, 1500 E. Medical Center Dr., 5316 Rogel Cancer Center, Ann Arbor, MI 48109-0940. Co-first authors (Josh N. Vo, Yi-Mi Wu).

## Abstract

Short-read sequencing cannot determine whether co-occurring variants within a cancer gene lie on the same allele (*cis*) or opposing alleles (*trans*), a distinction with direct therapeutic consequences: *trans* configurations confirm biallelic tumor suppressor inactivation, whereas *cis* configurations generate compound oncogenic alleles with enhanced activity. Among 768 patients with prostate, breast, or ovarian cancers, we used mutational signatures to nominate cryptic genomic instability cases lacking a causative biallelic event on short-read sequencing. Long-read nanopore sequencing resolved 32 of 46 cryptic cases (69.6%) through methylation detection, long insertion resolution, and structural variant characterization, confirming *trans* inactivation in every resolved tumor suppressor case. Analysis of 4,496 MiOncoSeq samples identified 17,519 multi-hit gene pairs, 78.7% of which exceeded the 500 bp short-read phasing limit, and long-read phasing revealed recurrent compound *cis* alleles in *NOTCH1*, *PIK3CA*, *PDGFRB*, and *KIT*. Haplotype phasing addresses an overlooked gap in cancer variant interpretation and warrants integration into precision oncology.

**Statement of Significance:** Short-read sequencing cannot resolve whether co-occurring variants within a cancer gene are *cis* or *trans*, a distinction critical for clinical interpretation. Long-read nanopore sequencing addresses this gap through haplotype phasing, methylation detection, and structural variant resolution, confirming biallelic tumor suppressor inactivation and revealing compound *cis* oncogenic alleles with enhanced activity.

## Introduction

The molecular characterization of cancer has undergone significant advancement due to the implementation of second-generation sequencing technologies, commonly referred to as short-read sequencing. This progress has paved the way for precision medicine approaches that align patients with targeted therapies based on the molecular profiles of their tumors.^1,2^ Nevertheless, conventional short-read sequencing platforms possess inherent limitations, particularly in their ability to detect complex structural variants, resolve repetitive genomic regions, and characterize long insertions that may contain clinically actionable information.^3,4^ These diagnostic shortcomings are further exacerbated by the use of targeted gene panels, which, while offering cost-effectiveness and rapid turnaround times, provide limited genomic coverage and resolution.^5^

In contrast, third-generation long-read nanopore sequencing effectively addresses these fundamental limitations. This technology facilitates the detection of complex structural variants and long insertions that are often overlooked by short-read methods, enables the characterization of repetitive elements, and supports haplotype phasing — the experimental resolution of which variants reside on the same versus opposing chromosomal alleles.^3,4,6^ Moreover, nanopore platforms can simultaneously detect epigenetic modifications, such as cytosine methylation, without the need for bisulfite treatment.^6^

Haplotype phasing carries distinct biological and therapeutic consequences depending on the gene context. When two variants co-occur within a cancer gene, their allelic configuration — whether on the same chromosome (*cis*) or on opposing alleles (*trans*) — determines their biological meaning entirely. For tumor suppressor genes, *trans* configuration confirms biallelic inactivation consistent with Knudson’s two-hit model, establishing pathogenicity and eligibility for pathway-targeted therapies such as PARP inhibitors in homologous recombination deficiency (HRD).^7,8^ For oncogenes, *cis* configuration generates compound alleles whose synergistic functional consequences are qualitatively distinct from either variant alone.^9^ Short-read sequencing cannot resolve phase for the majority of clinically relevant co-occurring variant pairs, and computational inference from variant allele frequencies is unreliable in the context of tumor heterogeneity and copy number alterations.^10^

In this study, we systematically evaluated the clinical utility of nanopore sequencing within a cohort of patients with advanced cancers exhibiting suspected genomic instability. First, we ascertain the incremental diagnostic yield of nanopore sequencing in identifying cryptic variants that conventional methods fail to detect and demonstrate that haplotype phasing experimentally confirms *trans* biallelic tumor suppressor inactivation underlying these cryptic genomic instability phenotypes. Second, through systematic analysis of 3,688 patients in the MiOncoSeq clinical sequencing program, we reveal a previously uncharacterized landscape of compound *cis* oncogenic alleles in *NOTCH1*, *PDGFRB*, *KIT*, and *PIK3CA*, with functional evidence of enhanced oncogenic activity and implications for targeted therapy selection.

## Results

### Systematic Identification of Cases with Cryptic Genomic Instability

We enrolled 768 patients with prostate, breast, or ovarian cancers in the PROBLEM cohort (<u>Pr</u>ostate-<u>O</u>varian-<u>Br</u>east-<u>L</u>ikely-<u>E</u>lusive-<u>M</u>utations) through the MiOncoSeq precision oncology program at the University of Michigan Rogel Cancer Center (**Table 1**, **Table S1**, and **Methods**). The cohort was assembled to test whether cases of genomic instability can be resolved to their causative biallelic driver, and was built on a phenotype-first principle: patients were selected by mutational-signature evidence of instability rather than by any predefined driver gene, identifying tumors that must carry a biallelic driver while remaining agnostic to its identity and avoiding the bias of looking only where a mutation is already known. Because these instability phenotypes arise from biallelic tumor suppressor inactivation, the expected driver is a two-hit configuration with the lesions on opposing alleles (in *trans*), making the cohort a natural setting to resolve such *trans* configurations directly. All patients underwent short-read whole-exome and targeted panel sequencing as part of routine clinical care, followed by computational mutational signature analysis to identify genomic instability phenotypes; cases lacking identifiable biallelic drivers despite strong signature evidence were nominated for long-read nanopore sequencing on the Oxford Nanopore PromethION platform (**Figure 1A-B**, **Figure S1**, and **Methods**), contingent on adequate DNA quality and quantity. Across this retrospective cohort, the interval from sample receipt to amended clinical report ranged from 5 to 30 days (mean 20, median 17), consistent with feasibility in a clinical reporting window (**Figure 1C**).

**Figure 1.**
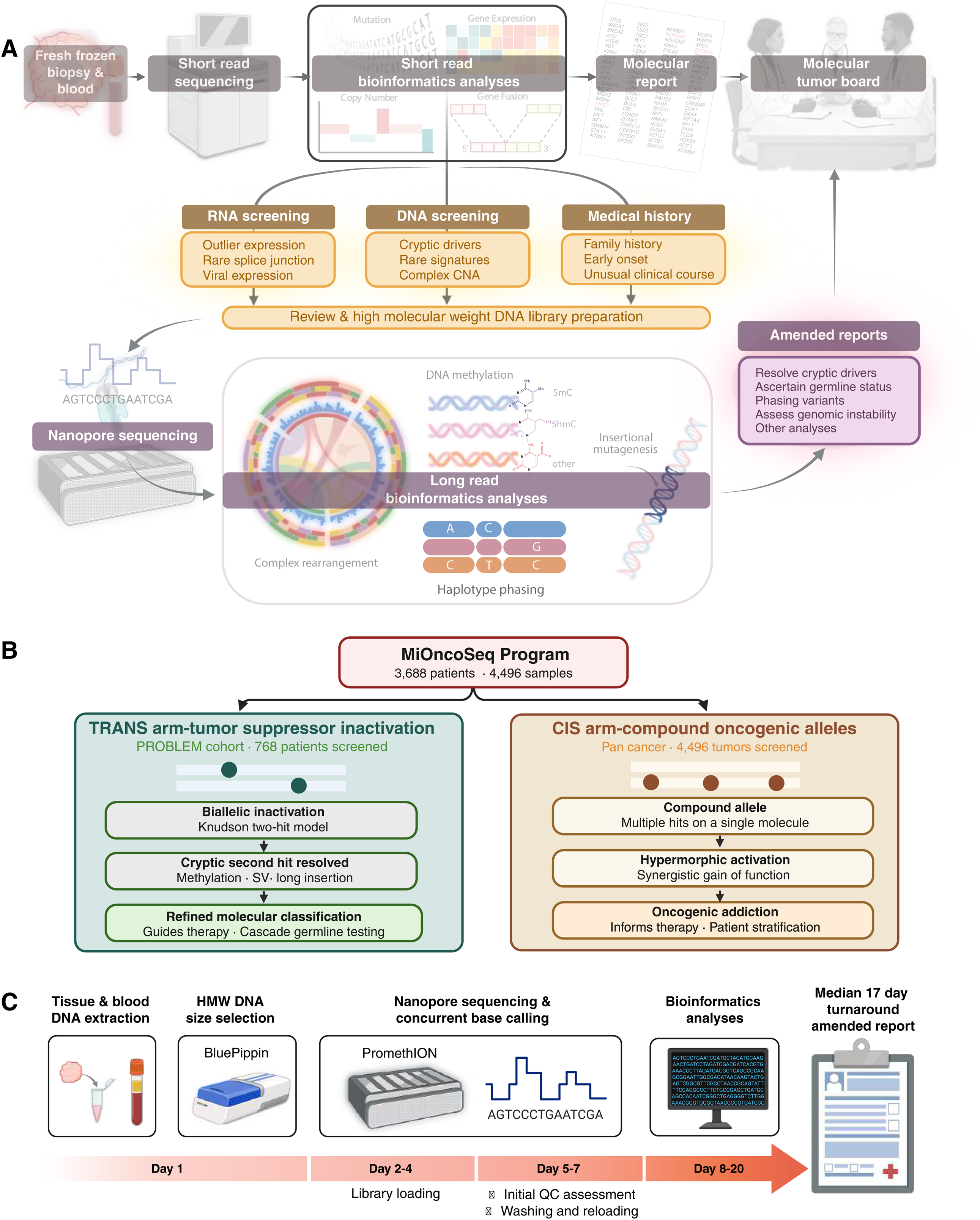
Overview of the integrative long-read sequencing workflow. **(A)** Clinical samples undergo standard short-read whole-exome and targeted panel sequencing as part of routine precision oncology care. Cases are nominated for long-read nanopore sequencing based on three complementary screening criteria: RNA evidence (outlier gene expression, rare splice junctions, viral transcript detection); DNA evidence (cryptic driver candidates, rare mutational signatures, complex copy number architecture); and relevant medical history (family history of cancer, early disease onset, unusual clinical course). Long-read bioinformatics analysis simultaneously characterizes complex structural rearrangements, performs haplotype phasing, and detects DNA methylation directly from base modification signals. Findings are communicated as amended clinical reports to treating oncologists. **(B)** Study design and analytical framework. Long-read sequencing was applied along two complementary axes. In the *trans* arm (tumor-suppressor inactivation), cases from the PROBLEM cohort (768 patients screened) were nominated by genomic-instability phenotype (homologous recombination deficiency, focal tandem duplication, and mismatch repair deficiency); haplotype phasing resolves whether two variants lie on opposing alleles (*in trans*), establishing biallelic inactivation, recovering cryptic second hits invisible to short-read sequencing (methylation, structural variants, long insertions), and refining molecular classification to guide therapy and cascade germline testing. In the *cis* arm (compound oncogenic alleles), recurrently multi-hit oncogenes across the pan-cancer MiOncoSeq cohort (4,496 tumors screened) were phased to determine whether co-occurring variants reside on the same allele (*in cis*), generating compound alleles with synergistic, hypermorphic gain of function that inform targeted therapy and patient stratification. **(C)** The timeline illustrates the representative workflow from sample receipt to amended report. Across the retrospective cohort, processing time ranged from 5 to 30 days (mean 20, median 17); the day ranges shown depict typical stage durations under batched research processing rather than a validated clinical turnaround. (A, Created in BioRender. Vo, J. (2026) https://BioRender.com/v21p25z)

**Table 1.** Patient Characteristics in the PROBLEM Cohort.

| **Table 1. Patient Characteristics in the PROBLEM Cohort.** |  |  |  |
| --- | --- | --- | --- |
| **Characteristic** | **Breast Cancer**<br>**(n=246)** | **Prostate Cancer**<br>**(n=505)** | **Ovarian Cancer**<br>**(n=17)** |
| **Cancer Subtype** |  |  |  |
| Ductal | 212 (86.2%) | — | — |
| Lobular | 24 (9.8%) | — | — |
| Other | 10 (4.1%) | — | — |
| mCRPC | — | 354 (70.1%) | — |
| Hormone naïve | — | 126 (25.0%) | — |
| Neuroendocrine | — | 25 (5.0%) | — |
| Serous | — | — | 14 (82.3%) |
| Non-serous | — | — | 3 (17.6%) |
| **Sex** |  |  |  |
| Male | 2 (0.8%) | 505 (100.0%) | 0 (0.0%) |
| Female | 244 (99.2%) | 0 (0.0%) | 17 (100.0%) |
| **Median age — yr (range)** | 56 (21-83) | 68 (36-93) | 62 (28-77) |

| **Race — no. (%)** |  |  |  |
| --- | --- | --- | --- |
| White | 221 (89.8%) | 435 (86.1%) | 15 (88.2%) |
| Black | 20 (8.1%) | 53 (10.5%) | 1 (5.9%) |
| Asian | 4 (1.6%) | 12 (2.4%) | 1 (5.9%) |
| Other | 1 (0.5%) | 5 (1.0%) | 0 (0.0%) |
| **Receptor status — no. (%)** |  |  |  |
| ER-positive, HER2-negative | 117 (47.6%) | — | — |
| HER2-positive | 23 (9.3%) | — | — |
| Triple negative | 106 (43.1%) | — | — |

Computational mutational signature analysis (**Methods**) identified 164 cases (21.3%) as *potential* genomic instability cases based on their signature profiles (**Figure 2A**): HRD (n=106 across prostate, breast, and ovarian cancers), focal tandem duplication (FTD; n=34, predominantly prostate), and mismatch repair deficiency (MMRD; n=24). Short-read sequencing immediately resolved a definitive biallelic driver in 118 of these 164 cases, leaving 46 cryptic cases that carried strong signature evidence of instability but no identifiable second hit; these 46 were selected for long-read nanopore sequencing (**Table S1, Figure 2A**). We additionally sequenced 14 positive-control cases (signature-positive tumors with a biallelic driver already resolved by short-read sequencing) and 53 negative-control cases (lacking any instability signature), giving a total of 113 PROBLEM-cohort tumors sequenced by long-read.

**Figure 2.**
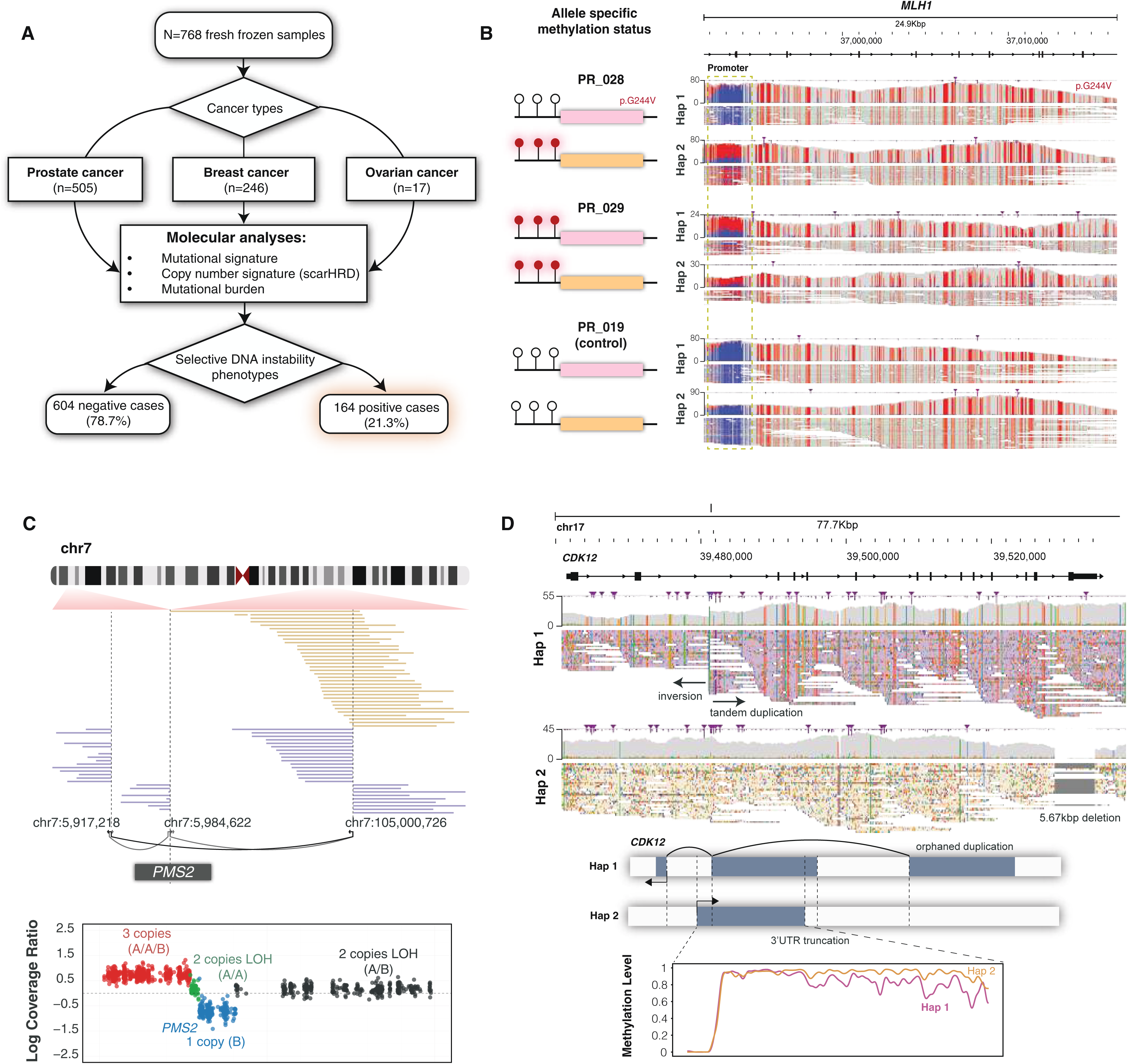
Cryptic MSI and FTD mechanisms resolved by long-read sequencing in the PROBLEM cohort. **(A)** Molecular diagnostic workflow for the PROBLEM cohort, showing computational mutational signature-guided nomination of 164 cases with genomic instability phenotypes from 768 patients with prostate, breast, or ovarian cancers. **(B)** Representative cryptic MMRD mechanism: allele-specific methylation tracks across the *MLH1* promoter demonstrating coordinated biallelic hypermethylation in two cases (PR_028, PR_029) relative to an unmethylated control (PR_019). **(C)** Long-read structural variant characterization of a chromothripsis-like *PMS2* rearrangement in a neuroendocrine prostate cancer case (PR_032). Short-read copy number analysis identified a hemizygous deletion of the *PMS2* region but could not resolve the second hit, which was characterized only by long-read sequencing. **(D)** Representative cryptic FTD mechanism: haplotype phasing enables detection of biallelic *CDK12* inactivation in PR_004. One allele harbors a 16 kb deletion removing the last exon and 3’ UTR, likely contributing to nonsense-mediated decay. The opposing allele contains an inversion truncating the first 20 kb of *CDK12* and a duplication of the orphaned segment. Haplotype-specific methylation analysis demonstrates progressive loss of epigenetic maintenance across the orphaned copies, with methylation levels declining toward the 3’ region.

### Long-Read Sequencing Resolves Cryptic Instability Through *Trans* Phasing

Cryptic MMRD mechanisms encompassed epigenetic and structural alterations, including coordinated *MLH1* promoter hypermethylation with biallelic transcriptional silencing, compound heterozygous *MLH1* methylation coupled with *trans*-acting mutation, and complex structural variants affecting *PMS2* and *MSH2* characterized by chromosomal rearrangements disrupting the remaining functional allele (**Figure 2B-C**, **Table S2**). Notably, the complex rearrangement involving *PMS2* in a patient with neuroendocrine prostate cancer exhibited chromothripsis-like, copy-neutral characteristics that evaded detection by short-read sequencing (**Figure 2C**).

FTD cases uniformly involved *CDK12* disruption through previously unrecognized balanced chromosomal rearrangements. Most of these reciprocal translocation events maintained copy number neutrality in one or both alleles, rendering them essentially invisible to conventional short-read targeted panel copy number analysis; yet, they were readily characterized through the capacity of long-read sequencing to traverse complex breakpoint junctions and resolve large-scale structural architecture (**Figure 2D**, **Figure S2**, **Table S3**).

Among HRD cases, *BRCA1* promoter hypermethylation with concurrent allelic loss emerged as the predominant mechanism in 10 of 20 resolved cases (50%) (**Figure 3A**, **Table S4**). Somatic *RAD51C* methylation was also observed in one patient (**Figure 3B**, **Table S4**). Cryptic *BRCA2* inactivation patterns were resolved in 4 cases, characterized by compound heterozygous deletions and a germline variant combined with structural rearrangements affecting the *trans* allele (**Table S4**). Focal *BARD1* deletions encompassing the critical RING domain were detected in 2 cases (one somatic, one germline), representing alterations that evade short-read detection due to the relative paucity of informative single-nucleotide polymorphisms (SNPs) in targeted panels (**Figure 3C**, **Table S4**). The somatic *BARD1* case harbored deletion of exons 2-3, which would produce isoform β (BARD1β) lacking the RING domain essential for BRCA1 binding, resulting in haploinsufficiency-mediated HRD through reduced functional BRCA1-BARD1 heterodimer formation.^11^ In the germline case, long-read-enabled haplotype phasing revealed an additional cryptic splicing variant of *BARD1* in *trans* with the germline deletion (**Figure 3C**, **Table S4**). A germline structural variant in *BRIP1* was observed in one patient with a history of breast and ovarian cancer (**Table S4**).

**Figure 3.**
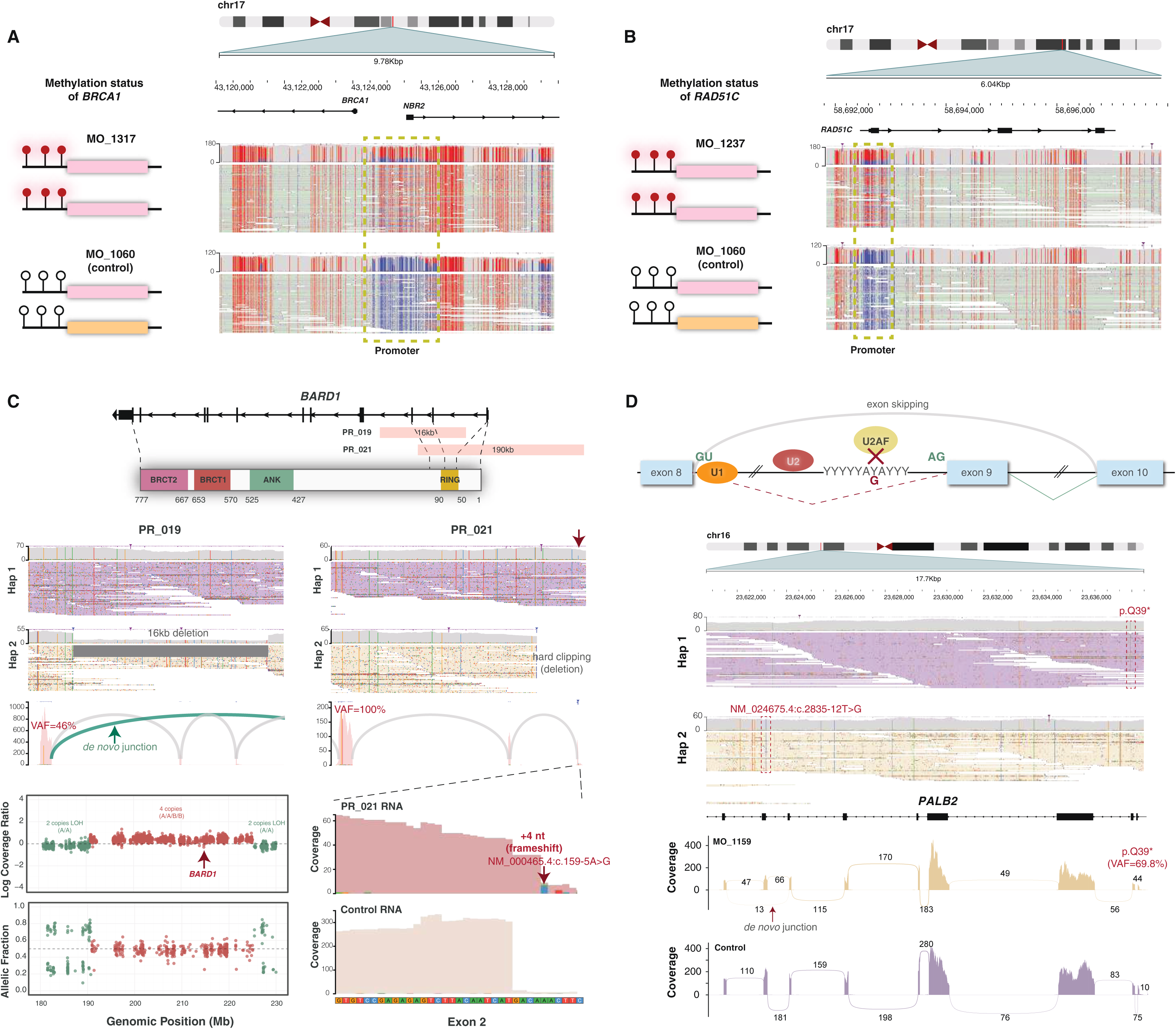
Cryptic HRD mechanisms resolved by long-read sequencing in the PROBLEM cohort. **(A) and (B)** Jbrowse screenshots show examples of promoter methylation affecting *BRCA1* and *RAD51C*. **(C) Top**: Gene diagram outlines two deletions affecting the RING domain of *BARD1* near the N- terminus. **Bottom**: Detailed genomic analysis. **Left**: The shorter deletion (16kb) is somatic, involving exons 2 and 3. This creates a *de novo* junction that corresponds to isoform beta (*BARD1*- β) that has been suggested to possess oncogenic activities. Interestingly, while this deletion is easily visible in JBrowse visualization of long-read sequencing, it escaped short-read sequencing detection due to two reasons: 1) the breakpoints are located in deep intronic regions, and 2) the paucity of SNPs in such a focal region evaded the segmentation methods used to detect copy number changes (bottom panel). **Right**: The longer deletion is germline (190kb), removing exon 1, exon 2, and the promoter. Integrative haplotype and allele-specific gene expression analysis demonstrated that expression derived entirely from the wild-type allele, ruling out alternative transcription initiation sites for the truncated *BARD1* variant. Careful inspection of the other allele revealed a somatic intronic variant upstream of the exon 2 splice acceptor (NM_000465.4:c.159- 5A>G) that creates a new splice acceptor, shifting the reading frame by 4bp. Thus, *BARD1* variants are biallelic in this case. **(D)** Patient PR_009 harbors NM_024675.4:c.2835-12T>G, an ultra-rare, previously unreported intronic *PALB2* splice variant. SpliceAI predicts a 94% probability of canonical splice acceptor loss through disruption of the polypyrimidine tract. **Top**: Schematic illustrating the proposed mechanism of exon 9 skipping resulting from impaired recognition of the 3′ splice acceptor site. **Middle**: Haplotype phasing demonstrates that the germline c.2835-12T>G variant occurs in *trans* with a somatic nonsense mutation (p.Gln39Ter), establishing biallelic *PALB2* inactivation. **Bottom**: RNA-sequencing analysis validates exon 9 skipping associated with the c.2835-12T>G variant.

An exemplary case involved a breast cancer patient in her 50s with an unremarkable family history exhibiting robust computational evidence for HRD yet harboring only a single somatic *PALB2* nonsense variant (p.Gln39Ter) by conventional analysis. Long-read sequencing uncovered an ultra-rare germline deep intronic splice variant positioned 12 base pairs upstream of the canonical splice acceptor site (NM_024675.4:c.2835-12T>G). Computational splice site prediction^12^ indicated the abolishment of normal splicing, which was experimentally validated through RNA sequencing, revealing the skipping of exon 9. Haplotype phasing definitively established the *trans* configuration of these variants, confirming biallelic *PALB2* inactivation and pathogenicity of the germline variant (**Figure 3D**, **Table S4**).

Initially, nanopore sequencing resolved 32 of 46 cryptic cases (69.6%), with complete diagnostic success for MMRD (5/5, 100%) and FTD (7/7, 100%) (**Table S2-S4**). In all but one *BARD1* case (which likely functions via a dominant-negative effect), biallelic inactivation was established by combining two loss-of-function events of distinct types, pairing a variant identified by conventional sequencing with a cryptic second hit that was a mutation, structural variant, or promoter methylation event. Haplotype phasing provided direct experimental confirmation that these events lay in *trans*, transforming a computationally inferred instability phenotype into an experimentally validated biallelic inactivation.

### Secondary Analyses of Unresolved Cases

The initial HRD resolution rate of 58.8% (20/34 cases) left 14 cases unresolved, prompting secondary signature analysis since computational signatures inferred from short-read sequencing may have lower specificity (**Methods**). Comprehensive mutational signature analysis using HRDetect^13,14^ scoring combined with detailed indel and structural variant characterization was performed using methodologies originally developed for short-read whole-genome sequencing but adapted for long-read data (more details in **Methods**). Specifically, we analyzed COSMIC indel signature 6 (ID6, deletions with microhomology) and indel signature 8 (ID8, deletions at repetitive sequences indicative of defective non-homologous end-joining repair), alongside structural variant signatures SV3 (short tandem duplications between 1-100Kb) and SV5 (short non-clustered deletions between 1-100Kb), which have been shown to be associated with HRD.^13–15^

Integrative analysis revealed that 11 of the 14 unresolved cases exhibited features inconsistent with classical HRD, including low HRDetect^13^ scores, isolated ID8 signatures without accompanying ID6 patterns, or complete absence of both ID6/ID8 and SV3/SV5 signatures despite initial computational HRD classification (**Figure 4A-B, Table S5** and more details in the **Methods**). Notable examples among these atypical cases included tandem duplication of *RAD51*’s initial exons, potentially altering expression of this essential gene.^16^ We also identified deep deletion of *ERCC4*, a nucleotide excision repair gene^17^ whose loss may force cellular reliance on homologous recombination pathways, overwhelming this repair system and producing HRD-like phenotypes. Only 3 of the 14 cases retained consistent HRD characteristics across all analytical dimensions (mutational patterns, copy number alterations, structural rearrangements, and indel signatures), yielding a *refined* diagnostic accuracy of 20/23 credible HRD cases (86.9%).

**Figure 4.**
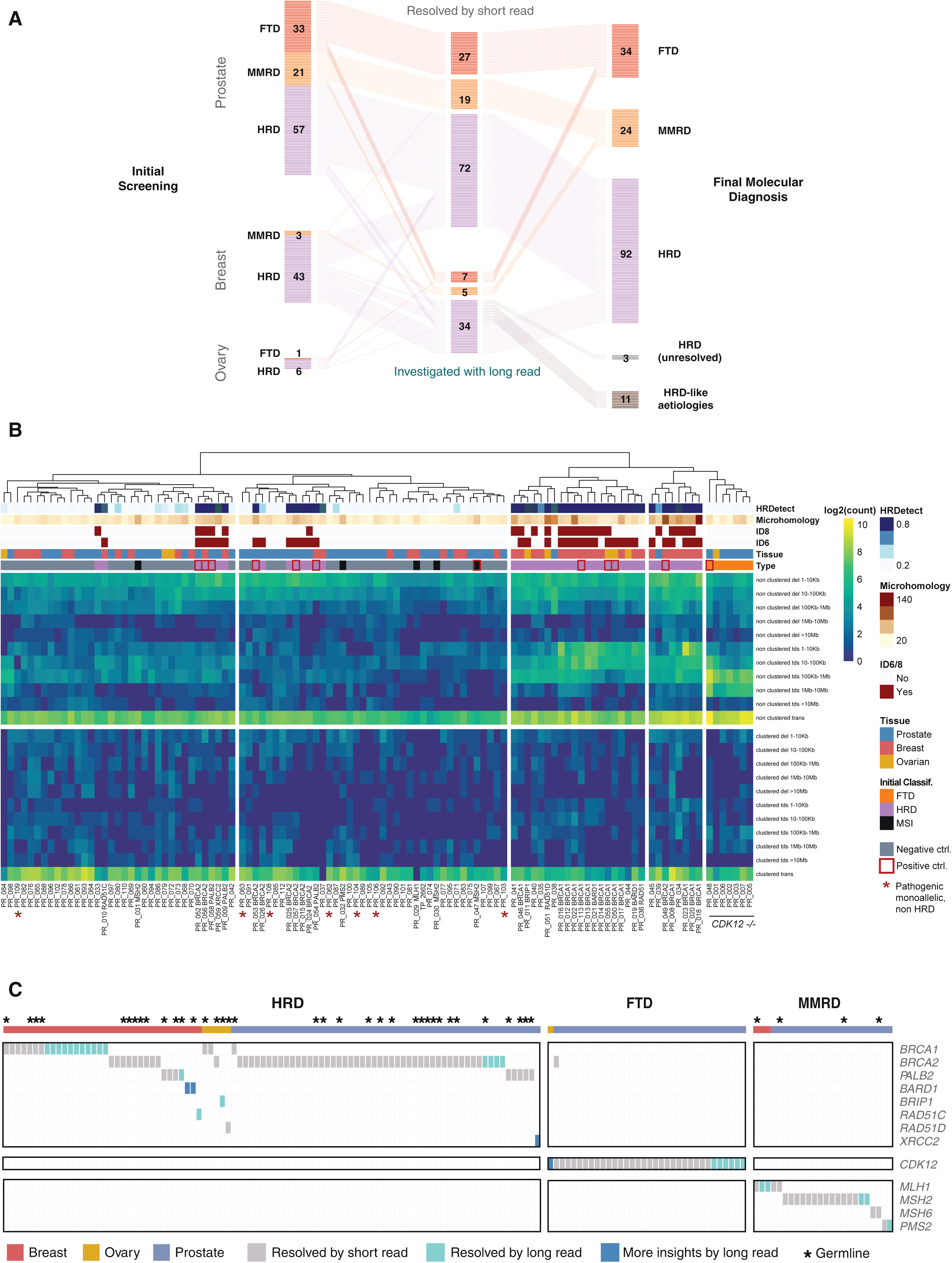
Diagnostic yield and molecular refinement of long-read sequencing in the PROBLEM cohort. **(A)** Alluvial diagram illustrating the flow of diagnostic resolution across MMRD, FTD, and HRD categories in prostate, breast, and ovarian cancers, from initial signature classification through short-read and long-read sequencing, including reclassification of 11 ambiguous HRD cases as HRD-like aetiologies following secondary signature analysis. **(B)** Heatmap displays structural variant features including insertions and deletions stratified by length, extracted using the *SigProfilerExtractor* package. *SigProfilerExtractor* categorizes structural variants as clustered or non-clustered; for this analysis, only non-clustered events were utilized to prevent artificial clustering bias from chromothripsis-associated "clustered" samples. Features underwent log2 transformation to normalize the wide range of structural variant counts and enable comparison across different event types. Manhattan distance calculation was employed because it is more robust to outliers and better suited for count-based data, particularly when analyzing discrete structural variant events. Complete linkage clustering methodology was applied to maximize between-cluster separation and create distinct, well-separated mutational pattern groups. Additional annotation tracks provide complementary mutational signature information: COSMIC indel signatures ID6 and ID8 (classified as positive when contribution ≥20%), quantification of microhomology sequences ≥3bp flanking structural variant breakpoints, and HRDetect scores for homologous recombination deficiency assessment. The analysis incorporates positive control cases with known driver alterations (indicated as red box) identified through short- read sequencing and negative control cases (indicated as gray box in annotation track) definitively lacking genomic instability signatures, providing validation benchmarks to guide interpretation of cryptic mutational mechanisms. Cases with pathogenic germline variants but lacking HRD phenotype were marked with red asterisks. For these analyses, tumor samples without matched normal controls were excluded. All FTD cases clustered together due to their characteristic tandem duplications ranging from 0.1 MB to 10 MB. Overall, breast cancer and ovarian cancer cases with HRD caused by *BRCA1*/*BARD1* alterations also clustered together, reflecting their enrichment for short tandem duplications and short deletions. Some HRD samples with *BRCA2* and *PALB2* deficiency showed less genomic scarring, characterized primarily by enrichment for short deletions. MMRD cases lacked characteristic copy number signatures, consistent with their primary manifestation through point mutations rather than structural alterations. **(C)** Oncoprint of gene-level mechanisms resolved across HRD, FTD, and MMRD cases, distinguishing events resolved by short-read sequencing (gray), resolved by long-read sequencing (teal), and providing additional mechanistic insight through long-read analysis (blue). Asterisks denote pathogenic germline variants.

Integrative secondary signature analyses also provided additional diagnostic and molecular insights. One prostate cancer patient harbored a homozygous deletion of *XRCC2* (which belongs to the RAD51 paralog complex BCDX2 with RAD51C, RAD51D, and RAD51B)^18^ initially detected by short-read sequencing. Given the rarity of *XRCC2* deficiency in clinical practice, integrative analyses were instrumental to confirm that this patient exhibited features of HRD, potentially benefiting from PARP inhibitor (PARPi) therapy (**Figure S3**). Conversely, 7 patients harbored known pathogenic germline variants or deleterious somatic variants (4 *BRCA2*, 2 *BRCA1*, 1 *BRIP1*) identified through short-read data review. Inspection of the *trans* alleles using long-read sequencing did not reveal additional hits, and comprehensive signature analyses reassured that these cases were non-HRD, indicating they would not benefit from PARPi therapy despite carrying pathogenic germline variants (**Figure 4B**).

Overall, long-read sequencing resolved 32 of 35 credible genomic instability cases (91.4%), and combined with short-read sequencing, the comprehensive diagnostic yield reached 91.4% for genomically unstable cancers (150 of 164 signature-positive cases) (**Figures 4A** and **4C**). Across these 150 resolved cases, the drivers segregated cleanly by phenotype: HRD (n=92) was driven predominantly by *BRCA2* (n=56) and *BRCA1* (n=21), with *PALB2* (n=9), *BARD1* (n=2), and single cases of *BRIP1*, *RAD51C*, *RAD51D*, and *XRCC2*; FTD (n=34) was uniformly driven by *CDK12*; and MMRD (n=24) by *MSH2* (n=15), *MLH1* (n=5), *MSH6* (n=2), and *PMS2* (n=2) (**Figure 4C**, **Table S1**). Notably, *BRCA1* inactivation showed a tissue-specific asymmetry, predominating in breast cancer (18 of 34 resolved HRD cases) but rare in prostate (1 of 53), consistent with its limited role in prostate cancer initiation and low germline prevalence in this setting.^19^ Long-read sequencing thus increased the diagnostic yield among signature-positive instability cases from 72.0% (118 of 164) to 91.4% (150 of 164), a gain of 19.5 percentage points, with the combined short- and long-read approach informing clinical management across the cohort (**Figures 4A-C**).

Long-read sequencing further demonstrated its capacity to resolve cryptic events beyond the instability-signature gene set, both in other somatic tumor suppressors (two *TP53* cases resolved by *trans* phasing and by deletion-plus-LOH; **Figure S4A-B**) and in the germline. Outside of the PROBLEM cohort, a woman in her 60s with a strong family history of breast and ovarian cancer had previously received negative results from commercial multi-gene panel testing and was evaluated outside our institutional cohort. Long-read nanopore sequencing of germline DNA from peripheral blood identified a pathogenic *AluYb8* retrotransposon insertion within exon 25 of *BRCA2* in reverse orientation. The inserted sequence exhibited 99.7% identity to the *AluYb8* consensus, differing by only a single base pair, consistent with a relatively recent insertion event in evolutionary terms.^20^ Target site duplications flanking the insertion were clearly resolved by long reads spanning the full insertion in single molecules, a structural feature invisible to short-read sequencing and absent from standard panel coverage (**Figure S5**). This case illustrates that long-read germline sequencing is warranted in patients with compelling personal or family histories and uninformative conventional testing, and that the diagnostic gap extends beyond computational instability signatures.

### The Scope of Co-Occurring Variants Requiring Phase Determination

Having established that *trans* phasing is essential for confirming tumor suppressor biallelic inactivation, we asked how broadly the phase ambiguity problem extends across the clinical sequencing landscape. Prior pan-cancer analyses have demonstrated that compound mutations within individual cancer genes are a common phenomenon: among significantly enriched oncogenes, up to 9% of mutated samples harbor co-occurring mutations,^9^ while across all cancer-associated genes, nearly one in four advanced tumors contain composite mutations.^21^ In both studies, oncogene compound mutations were predominantly *cis*-acting, with 78-91% residing on the same allele, yet these estimates relied on RNA sequencing reads or short-read WES data, approaches inherently restricted to variants within a single read length and unable to resolve the majority of clinically relevant pairs.^9,21^ Systematic analysis of 4,496 MiOncoSeq samples across 3,688 patients (**Figure 1B**) identified 17,519 co-occurring variant pairs within individual cancer genes, confirming the prevalence of this phenomenon in a clinical sequencing cohort. Multi-hit events were prevalent across both tumor suppressors and oncogenes: among tumor suppressors, *TET2* (23.9%), *NOTCH1* (22.9%), and *ASXL3* (22.4%) showed the highest multi-hit frequencies, while oncogenes including *MYC* (21.3%), *FGFR3* (16.4%), *KIT* (15.8%), *PIK3CA* (12.9%), were also frequently affected (**Figure 5A**). Whether these co-occurring oncogene variants reside on the same allele, generating hypermorphic configurations with qualitatively distinct oncogenic properties, has remained experimentally unresolvable at clinical scale.

**Figure 5.**
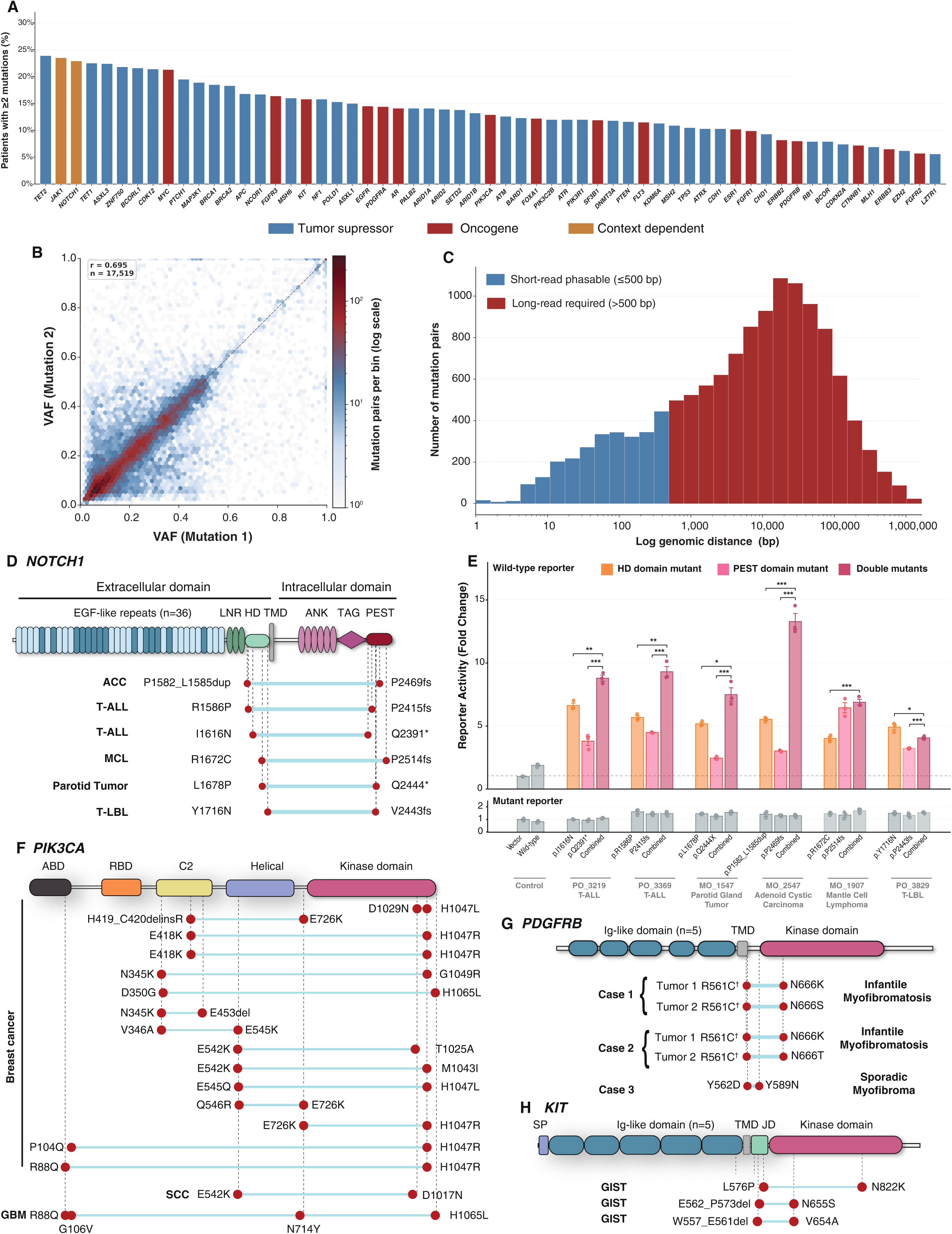
Compound *cis* oncogenic alleles across cancer types revealed by long-read haplotype phasing. **(A)** Frequency of multi-hit events across curated cancer genes in 4,496 tumor samples (3,688 patients). Bars represent the percentage of samples with ≥2 somatic mutations among all samples harboring any mutation in that gene, ordered by decreasing frequency. Blue, tumor suppressor; red, oncogene; amber, context-dependent. **(B)** VAF correlation of co-occurring SNV pairs within the same gene and sample (n = 12,934). Hexagonal bins colored by log-scaled pair count; white bins indicate zero observations. Dashed line, VAF₁ = VAF₂. Pearson r reported. **(C)** Genomic distance distribution of co-occurring SNV pairs (n = 12,934; log-scaled x-axis). Blue, phasable by short-read sequencing (≤500 bp); red, requiring long-read sequencing (>500 bp). **(D)** NOTCH1 protein schematic showing compound *cis* variant pairs (teal bars) confirmed by long-read phasing across six patients, consistently pairing heterodimerization (HD) domain with PEST domain variants. **(E)** Luciferase reporter assays measuring transcriptional activation of compound NOTCH1 HD+PEST mutations relative to individual variants. Four of six compound pairs showed significantly enhanced NOTCH pathway activation. Unpaired two-tailed Welch’s t-test; n = 3 per group; *p < 0.05, **p < 0.01, ***p < 0.001. **(F)** PIK3CA protein schematic (ABD, RBD, C2, helical, kinase domains) showing compound *cis* variant pairs confirmed by long-read phasing in breast cancer and additional tumor types. Each row represents one patient; circles denote variant positions; dashed lines indicate confirmed *cis* configuration. **(G)** PDGFRB protein schematic showing compound *cis* variants in three mesenchymal tumors: two infantile myofibromatosis cases with germline(†) p.Arg561Cys paired with somatic Asn666 alterations, and one sporadic myofibroma with dual somatic tyrosine substitutions. **(H)** KIT protein schematic showing compound *cis* variants across three cases, spanning juxtamembrane and activation loop domains.

Variant allelic fraction (VAF) correlation analysis restricted to SNV pairs (n=12,934) revealed strong diagonal clustering (r=0.695), a pattern consistent with, but not diagnostic of, *cis* configuration (**Figure 5B**). This ambiguity is precisely the clinical problem: correlated VAFs can reflect *cis* compound alleles or clonal co-evolution of *trans* variants and cannot be distinguished without experimental phasing. Analysis of genomic distances demonstrated that 78.7% of pairs spanned distances exceeding 500 bp, the practical phasing limit of short-read sequencing, with a median inter-variant distance of 8,420 bp (**Figure 5C**). Notably, a substantial fraction of pairs exceeded 100,000 bp, driven by mutations in exceptionally large genes such as *LRP1B* (∼1.9 Mb) and *NF1* (∼280 kb), distances that demand ultra-long nanopore reads for resolution.

The same phase ambiguity extends to germline-somatic configurations. Across 120 curated hereditary cancer genes in the matched cohort, we identified 41 germline-somatic variant pairs across 39 patients and 27 genes, including two-hit events in *DICER1*, *APC*, *TP53*, *NF1*, *MLH1*, *SMARCA4*, and *PDGFRB* (**Methods**). As with the somatic pairs, the vast majority (92.7%) exceeded the 500 bp short-read phasing limit (median 17,080 bp), confirming that germline-somatic compound configurations are likewise unresolvable by short-read sequencing. The overwhelming majority of co-occurring variant pairs in the clinical sequencing landscape, whether somatic or germline-somatic, therefore require long-read sequencing for definitive allelic assignment, which we applied to systematically characterize compound *cis* alleles across recurrently multi-hit oncogenes.

### Compound Cis *NOTCH1* Variants Create Hypermorphic Oncogenic Alleles

NOTCH1 signaling is regulated through a precise autoinhibitory architecture in which the heterodimerization (HD) domain restrains the receptor in a quiescent state until ligand-induced cleavage, while the PEST domain governs intracellular domain degradation.^22^ In our cohort, 22.9% of *NOTCH1*-mutated samples harbored co-occurring variants (**Figure 5A**), reflecting its context-dependent role as both an oncogene and tumor suppressor across different malignancies.^9,21,22^ Among these, a consistent pattern emerged: a subset of patients harbored one variant in the HD domain paired with one in the PEST domain. Six patients with available high-molecular-weight DNA were selected for long-read sequencing. Long-read sequencing confirmed that, in every case, these variants resided in *cis* on the same allele, across a spectrum of diagnoses including T-cell acute lymphoblastic leukemia, mantle cell lymphoma, adenoid cystic carcinoma, and parotid gland tumor (**Figure 5D**, **Figure S6**).

Functional validation using a *NOTCH1* transcription reporter assay demonstrated significantly greater target gene activation in four of six compound HD+PEST double mutants compared to either single mutant alone (*p*<0.001 for significant pairwise comparisons), consistent with synergistic disruption of both autoinhibitory and degradation mechanisms (**Figure 5E**). The compound mutants showed reporter activity exceeding the additive effects of the individual variants, establishing that *cis* compound *NOTCH1* alleles constitute a qualitatively distinct oncogenic entity. To further determine whether this synergy extends beyond reporter activity to the broader transcriptional program, we performed RNA-seq on the same system, which confirmed that the compound allele intensifies the canonical NOTCH1 target program in a dose-dependent manner while also eliciting non-additive, synergistic changes in additional pathways, consistent with a qualitatively distinct expression program (**Figure S7A-C**). Whether these compound alleles exhibit altered sensitivity to *NOTCH1*-targeted agents or exhibit poorer survival warrants prospective investigation.

### Compound *Cis PIK3CA* Alleles Are Prevalent and Arise in Recurrent Domain-Specific Configurations

*PIK3CA* activating mutations occur in approximately 40% of hormone receptor-positive breast cancers and form the basis for PI3Kα inhibitor therapy with alpelisib, approved in combination with fulvestrant for this indication.^23,24^ The functional consequences of co-occurring *PIK3CA* mutations have been established by complementary pan-cancer studies demonstrating that compound mutations within *PIK3CA* arise more frequently than expected by chance, preferentially in *cis*, and confer enhanced oncogenic activity beyond either variant alone.^9,21,25^ However, the allelic configuration of these variants and therefore their functional and therapeutic interpretation could not be experimentally confirmed by short-read sequencing in prior work.

Among samples with HMW, long-read sequencing identified compound cis *PIK3CA* alleles in 15 breast cancer patients as well as cases spanning brain and squamous cell carcinoma (**Figure 5F**). Variant pairs encompassed combinations across all five structural domains of p110α — the adaptor-binding domain (ABD), Ras-binding domain (RBD), C2 domain, helical domain, and kinase domain — indicating that compound *PIK3CA* alleles are not restricted to specific hotspot combinations but arise across the full mutational landscape of the gene. The combination spanning the greatest genomic interval in our breast cancer cohort involved p.R88Q in the ABD and p.H1047R in the kinase domain, separated by approximately 35.2 kilobases (chr3:178,916,876 to chr3:178,952,085) (**Figure 5F**), a distance entirely beyond short-read phasing resolution and confirmed in *cis* only through long-read sequencing.

Two cases beyond breast cancer illustrate the clinical breadth and biological significance of compound *PIK3CA* alleles. A pediatric patient with radiation-induced brainstem glioblastoma harbored four concurrent *PIK3CA* variants confirmed in *cis* by nanopore sequencing: p.R88Q and p.G106V in the ABD, and p.N714Y and p.H1065L in the kinase domain. Despite this extreme allelic complexity, the overall tumor mutation burden was low, indicating that these four variants did not accumulate passively but were each subject to strong positive selection. This pattern is consistent with oncogenic addiction^26^ to a massively hypermorphic *PIK3CA* allele in which progressive intramolecular destabilization across multiple domains drives pathway dependence beyond what any single mutation could achieve.

In contrast, a patient with ocular squamous cell carcinoma, HPV-positive and hypermutated through APOBEC-associated mutagenesis,^27^ harbored three *PIK3CA* mutations: p.E542K in the helical domain, p.R93W in the ABD, and p.D1017N in the kinase domain. Long-read phasing revealed that only p.E542K and p.D1017N resided in *cis*, while p.R93W occupied the opposing allele in *trans*. Critically, all three variants were present at nearly identical variant allele frequencies of approximately 15%, a finding that would conventionally suggest the variants are clonal and indistinguishable in their allelic relationships. This case directly illustrates that VAF is nondiagnostic for phasing in the context of hypermutation and complex clonal architecture, and that only experimental long-read phasing can resolve the true allelic configuration.

### Compound *Cis* Variants in Mesenchymal Tumors

*PDGFRB* and *KIT* are paralogous members of the Type III receptor tyrosine kinase family, sharing five extracellular immunoglobulin-like domains, a juxtamembrane autoinhibitory segment, and a split kinase domain.^28^ Both drive distinct mesenchymal tumor types and are therapeutically targeted by imatinib, making allelic resolution of compound variants in these genes directly actionable.^29,30^

*PDGFRB* kinase activity is regulated through juxtamembrane autoinhibitory interactions involving key residues including Arg561, and germline p.Arg561Cys is the canonical pathogenic variant in familial infantile myofibromatosis.^28,29^ Long-read sequencing confirmed that p.Arg561Cys was in compound *cis* with somatic Asn666 alterations in two independent cases of infantile myofibromatosis, providing the first direct experimental allelic confirmation of the known two-hit mechanism in this disease (**Figure 5G**, **Figure S6**). Interestingly, a third sporadic myofibroma case harbored a novel compound somatic configuration: p.Tyr562Asp paired with p.Tyr589Asn in *cis*, representing the first description of dual tyrosine alterations affecting both the juxtamembrane domain and kinase domain N-lobe simultaneously (**Figure 5G**). While p.Arg561Cys disrupts a single critical autoinhibitory contact, this novel combination potentially destabilizes the juxtamembrane-kinase interface through two independent structural perturbations, raising the possibility of distinct sensitivity to tyrosine kinase inhibitors targeting this regulatory interface.

In gastrointestinal stromal tumor (GIST), primary activating *KIT* juxtamembrane mutations initiate oncogenic signaling, whereas acquired secondary kinase-domain mutations affecting either the ATP-binding pocket or activation loop represent the dominant mechanism of imatinib resistance.^30,31^ Prior mutational analyses suggested that these resistance mutations arise in *cis* with the primary driver allele, but direct experimental confirmation across kilobase-scale genomic intervals separating these variants has remained impractical with short-read sequencing.^32^ Long-read phasing in three patients with documented imatinib progression confirmed *cis* compound *KIT* alleles spanning three mechanistically distinct resistance architectures (**Figure 5H**): one activation-loop secondary mutation (p.N822K, associated with relative susceptibility to regorafenib-class inhibitors), and two ATP-binding-pocket mutations (the novel p.N655S and p.V654A, the latter classically associated with sunitinib sensitivity).^32^ Phase-aware genotyping in this setting directly informs rational tyrosine kinase inhibitor sequencing because the secondary mutation present on the primary driver allele predicts differential inhibitor susceptibility. In practice, confirming that a p.V654A or p.N822K secondary mutation resides on the primary driver allele identifies it as the actionable resistance target within the dominant clone, guiding selection of the inhibitor class active against that specific compound configuration (for example, sunitinib for ATP-binding-pocket and regorafenib for activation-loop mutations) rather than treating the two variants as independent events.^32^

Overall, long-read phasing confirmed a compound *cis* configuration in all *NOTCH1*, *PDGFRB*, and *KIT* cases (15/15) and in 16 of 17 *PIK3CA* cases, the exception being a single tumor with a co-occurring *trans* variant. Domain localization and variant allele frequency may guide prioritization of candidate compound *cis* alleles from short-read data, but only haplotype phasing by long-read sequencing establishes the configuration definitively.

## Discussion

Preliminary attempts have been made in recent years to integrate long-read sequencing into clinical settings, particularly for genetic disorders and select oncological applications.^33–35^ However, these efforts have largely been limited to highly specialized cases, with systematic integration into routine precision oncology workflows remaining largely unexplored. Our study represents one of the first comprehensive evaluations of long-read sequencing in a clinical precision oncology program and uniquely addresses two dimensions that have received insufficient attention: the detection of cryptic pathogenic variants invisible to conventional sequencing and the experimental resolution of variant phase across clinically relevant genomic distances.

While targeted short-read sequencing resolved 71.9% of cases with genomic instability signatures, it left 28.0% mechanistically unexplained despite strong computational evidence. Long-read sequencing resolved 69.6% of these previously cryptic cases, achieving complete resolution for all focal tandem duplications and mismatch repair deficiency cases, and resolving the majority of credible homologous recombination deficiency cases. The combined diagnostic approach yielded actionable findings that influenced treatment decisions in 19.5% of all patients studied. These findings support a **tiered** diagnostic algorithm that deploys long-read sequencing in contexts where it adds maximal value: tumors with strong genomic instability signatures but no identifiable biallelic driver; patients with compelling family histories or multiple primaries despite negative panel results; and tumors with atypical molecular or phenotypic features such as unexplained gene overexpression or paradoxical clinical behavior. As platform costs decline and analytical pipelines mature, we anticipate progressive movement of long-read sequencing toward earlier implementation.

Beyond resolving co-occurring variants with known mechanisms, long-read sequencing uniquely detects pathogenic events that are structurally invisible to short-read methods — including complex balanced rearrangements maintaining copy number neutrality, deep intronic splice variants beyond panel coverage, and mobile element insertions such as the *AluYb8* retrotransposon identified in our germline *BRCA2* case. Crucially, methylation is detected simultaneously from the same nanopore run without bisulfite treatment, enabling direct epigenetic characterization of promoter silencing and allele-specific imprinting in the same assay that performs variant calling and haplotype phasing.

The haplotype phasing capabilities of long-read sequencing translate directly into clinical decision-making. When a patient with a mismatch repair deficiency signature but no identifiable biallelic driver is shown to carry biallelic somatic *MLH1* promoter methylation, this reduces the likelihood of an occult germline variant and the urgency of cascade family screening; conversely, demonstrating that a germline variant of uncertain significance lies *in trans* with a somatic second hit provides strong evidence for pathogenicity and directs counseling, as in our *PALB2* case, where a cryptic deep intronic splice variant would otherwise have gone undetected. Phasing equally resolves the inverse dilemma of apparently monoallelic inactivation: a single *BRCA1* mutation or hemizygous deletion with an HRD-suggestive signature may reflect either a cryptic second hit or a phenocopy, and long-read sequencing distinguishes these by combining structural, copy-number, methylation, and signature assessment with direct interrogation of the *trans* allele. The same caution applies in reverse: a germline carrier in *BRCA1*, *BRCA2*, *PALB2*, *BARD1*, *RAD51C*, or *BRIP1* should not be assumed to have functional HRD without confirmed biallelic inactivation. In every case, germline or somatic status alone is insufficient, and allelic phase supplies the missing evidence.

In the case of oncogenes, compound *cis* alleles in *EGFR*, *NOTCH1*, *PIK3CA*, *PDGFRB*, and *KIT* warrant systematic investigation to enable genotype-based patient stratification. It has long been proposed that tumors develop increased dependency on individual signaling pathways through oncogene addiction.^26^ The progressive accumulation of compound *cis* variants within a single oncogene represents a molecular substrate for this phenomenon, pointing to pathway-specific therapeutic vulnerabilities that may be invisible to gene-level mutation calls alone. Distinct compound allele combinations likely confer characteristic therapeutic responses and clinical outcomes not captured by single-variant classification.

A critical and underappreciated consequence of phase-agnostic clinical practice is that most trials enrolling patients on the basis of gene mutation status are blind to allelic configuration. As a result, trials may inadvertently mix monoallelic carriers and true compound mutant cases within the same enrollment arm, diluting effect sizes, obscuring genotype-response relationships, and potentially misattributing resistance or exceptional responses to the wrong molecular substrate. Prospective stratification by confirmed allelic phase is now technically feasible and should be incorporated into trial design going forward.

Current clinical genomics reporting frameworks do not routinely operationalize variant phase or epigenetic allele status as structured report elements. The Association for Molecular Pathology / American Society of Clinical Oncology / College of American Pathologists somatic variant interpretation guidelines emphasize genomic coordinates, transcript accession, variant allele fraction, and tier classification, but do not include allelic phase as a required field.^36^ The European Society for Medical Oncology 2024^37^ clinical genomic reporting recommendations likewise address major genomic biomarkers without defining how co-occurring variants within the same gene should be phased in clinical reports. By contrast, Human Genome Variation Society nomenclature formally supports phase annotation through bracket syntax: c.[2376G>C;3103del] denotes *cis* configuration, whereas c.[2376G>C];[3103del] denotes *trans*, yet this notation is rarely used in oncology reporting.

We propose that phase annotation and allele-specific methylation status be incorporated whenever multiple variants occur within the same clinically relevant cancer gene. Analogous to existing tiered reporting logic, a practical framework could distinguish **Level I**, phase experimentally confirmed by long-read or another long-range method; **Level II**, high-confidence inferred phase from VAF concordance or linkage; and **Level III**, phase explicitly reported as unresolved. Such determination is most warranted when co-occurring variants fall in HRD-, FTD-, or MMRD-associated genes with strong signature evidence, in oncogenes with known compound-allele biology, or when a variant of uncertain significance accompanies a known pathogenic variant in a tumor suppressor. Incorporating these fields would convert currently latent allelic information into structured, clinically interpretable annotations.

Allelic configuration and cryptic driver detection are two faces of the same interpretive gap: short-read sequencing neither resolves whether co-occurring variants inactivate a tumor suppressor or build a compound oncogenic allele, nor reliably detects the structural, epigenetic, and insertional mutagenesis that constitute the driver in the first place. Long-read sequencing addresses both in a single assay, converting variants that are currently invisible or ambiguous into resolved, clinically interpretable findings. Together these capabilities establish allelic phase and cryptic-variant resolution as necessary and now technically achievable dimensions of precision oncology.

## Methods

### Study Design and Patient Cohorts

This study undertook a retrospective analysis of patients who participated in the MiOncoSeq precision oncology program at the University of Michigan Rogel Cancer Center between January 2010 and December 2024. Patient samples, including fresh-frozen tumor tissue, matched blood, and buccal DNA, were collected following written informed consent under protocols approved by the University of Michigan Institutional Review Board (Michigan Oncology Sequencing Protocol, MiOncoSeq and PEDS_MiOncoSeq; IRB HUM00046018, HUM00067928, HUM00056496). All clinical sequencing was performed within our Clinical Laboratory Improvement Amendments (CLIA)-certified molecular diagnostics laboratory.

The study comprised two complementary screens that nominated cases for long-read sequencing. The first, the PROBLEM cohort (Prostate-Ovarian-Breast-Likely-Elusive-Mutations), comprised patients with advanced prostate cancer (n=505), breast cancer (n=246), and ovarian cancer (n=17) who had accessible frozen tumor tissue with greater than 30% tumor purity confirmed by pathological review (768 patients total; detailed demographics in Table 1). This cohort was ascertained by phenotype: cases were nominated on the basis of mutational-signature evidence of genomic instability rather than by any predefined driver gene. Of the 768 cases, 164 were signature-positive for genomic instability, of which 118 had a biallelic driver resolved by short-read sequencing, leaving 46 cryptic cases that were selected for long-read nanopore sequencing. To validate the long-read pipeline, we additionally sequenced 14 positive-control cases (signature-positive tumors with a biallelic driver already resolved by short-read sequencing) and 53 negative- control cases (lacking an instability signature), yielding 113 PROBLEM-cohort tumors sequenced by long-read.

The second screen drew on the broader MiOncoSeq cohort (n=4,496 samples from 3,688 patients),^38–40^ encompassing all enrolled cancer types, which was analyzed for systematic characterization of co-occurring variant pairs within individual cancer genes. From this screen, 32 tumors (30 patients) with recurrent compound-variant configurations in *NOTCH1*, *PIK3CA*, *PDGFRB*, and *KIT* were selected for long-read phasing to resolve their cis versus trans allelic configuration.

In total, 145 tumors underwent long-read sequencing across the two screens. One additional case, a germline *BRCA2* AluYb8 insertion identified outside the PROBLEM cohort, was analyzed by long-read sequencing of a normal sample to resolve the cryptic germline variant. Matched normals were sequenced where available; three PROBLEM-cohort matched normals derived from buccal samples failed library preparation and were excluded, leaving 43 matched normals for the 46 cryptic PROBLEM tumors.

### Integrative Clinical Sequencing by the Illumina Platform

Tumor genomic DNA and total RNA were extracted from fresh-frozen tissue using the AllPrep DNA/RNA/miRNA Kit (Qiagen). Matched normal genomic DNA was isolated from peripheral blood or buccal swab samples using the DNeasy Blood & Tissue Kit (Qiagen). For nanopore sequencing, DNA samples were size selected using Ampure XP beads in a buffer containing 0.75M NaCl and 20% PEG8000 to remove fragments smaller than 2 kb before library preparation. All cancer patients included in this study had previously undergone short-read sequencing on the Illumina platform. This included whole-exome sequencing, targeted exon sequencing, and/or capture-based RNA sequencing, as previously described.^38–40^

### Long-Read Sequencing Protocols

The size distribution of input genomic DNA was assessed using pulsed-field gel electrophoresis (Pippin Pulse; Sage Science). Samples with a high proportion of short DNA fragments were further processed using the BluePippin High-Pass DNA Size Selection System (Sage Science) to enrich for fragments greater than 5 kb, suitable for nanopore sequencing.

Sequencing libraries were prepared using the Ligation Sequencing Kit V14 (SQK-LSK114; Oxford Nanopore Technologies) in combination with the NEBNext Companion Module for Oxford Nanopore Technologies Ligation Sequencing (E7180S; New England Biolabs), following the manufacturers’ protocols. Briefly, 1 μg of high–molecular–weight (HMW) genomic DNA underwent end-repair, A-tailing, and adaptor ligation. Final libraries were purified using AMPure XP beads, quantified with a Qubit 3.0 Fluorometer (Thermo Fisher Scientific), and sequenced on the PromethION P24 platform equipped with A100 flow cells (Oxford Nanopore Technologies).

Of the compound-variant cases (**Table S1**), 17 were sequenced using adaptive sampling (Oxford Nanopore Technologies), which enriches for regions of interest by real-time selective rejection of off-target reads. The adaptive sampling panel targeted approximately 200 key tumor suppressor genes and oncogenes, each extended to include 100 kb upstream and downstream of the gene body to capture regulatory regions and distal structural variants.

### Cell lines

HEK293FT (RRID:CVCL_6911) cells were obtained from Invitrogen (Thermo Fisher Scientific, R70007) in 2013. Cell line authentication was performed by short tandem repeat (STR) profiling through Laboratory Corporation of America (Labcorp); STR profiles were compared with reference profiles in the Cellosaurus database using the CLASTR search tool or, when necessary, manually against reference profiles from public repositories (e.g., ATCC and DSMZ), with a cell line considered authenticated at ≥80% concordance. Cells were tested for *Mycoplasma* contamination biweekly using the MycoAlert PLUS Detection Kit (Lonza, LT07-710). All experiments used cells within 20 passages of thawing.

### NOTCH1 Reporter Assay

NOTCH1 reporter assays were conducted to assess the functional impact of individual and combined *NOTCH1* mutations. Constructs encoding single or double *NOTCH1* mutations were generated and cloned into mammalian expression vectors. HEK293FT cells were co-transfected with the *NOTCH1* mutant constructs, a NOTCH1-responsive reporter plasmid (RBP-Jk Reporter; SABiosciences/Qiagen), and a Renilla luciferase control plasmid (pRL-TK) using FuGENE HD (Promega), following the manufacturer’s protocol.

Forty-eight hours post-transfection, luciferase activity was measured using a dual-luciferase reporter assay system (Promega). Firefly luciferase activity, driven by the *NOTCH1*-responsive promoter, was normalized to Renilla luciferase activity to control for transfection efficiency. Fold activation of NOTCH signaling was calculated relative to cells transfected with empty vectors, enabling direct comparison of pathway activity across different mutant constructs.

### Short-Read Sequencing Bioinformatics Analyses

Whole-exome and targeted panel sequencing were performed on Illumina HiSeq and NovaSeq 6000 platforms, generating paired-end reads of 100-150 bp with average library fragment sizes of 350 bp. Raw sequencing reads underwent quality control using *FastQC* (RRID:SCR_014583). Alignment to the human reference genome (GRCh37/hg19) was performed using *NovoAlign* (v4.02.02, RRID:SCR_014818) with default parameters optimized for clinical sequencing. Post-alignment processing included duplicate marking and sorting using *NovoSort* (RRID:SCR_014818), followed by recalibration of base quality scores and local realignment around indels using GATK best practices.^41^

Somatic and germline variant calling employed a multi-caller approach utilizing *FreeBayes* (v1.3.6, RRID:SCR_010761) for single-nucleotide variants and small indels, and *Pindel* (v0.2.5 b8, RRID:SCR_000560)^42^ for structural variants and larger indels. Copy number alterations were detected using *cnatools*^38–40^ that jointly estimate tumor purity and ploidy. For RNA sequencing, reads were aligned using *STAR* (v2.7.10a, RRID:SCR_004463)^43^ with two-pass mode, and gene fusion detection was performed using *Arriba* (v2.4.0, RRID:SCR_025854)^44^ and *STAR-Fusion* (v1.15.0, RRID:SCR_025853) with stringent filtering criteria to minimize false positives. Ancestry (**Table 1**) was inferred from genotype data using *PLINK* (v1.9, RRID:SCR_001757)^45^ for quality control and linkage disequilibrium pruning, followed by *ADMIXTURE* (v1.3, RRID:SCR_001263)^46^ for model-based clustering analysis with reference populations from the 1000 Genomes Project.^47^

### Long-Read Sequencing Bioinformatics Analyses

Sequencing was performed on PromethION flow cells (R10.4.1 chemistry) with real-time basecalling using *Dorado* (RRID:SCR_025883) high-accuracy (hac) mode (Model: dna_r10.4.1_e8.2_400bps_hac@v5.0.0). Quality control assessment was conducted using *ToulligQC* (v2.7.1, RRID:SCR_028846), which provided comprehensive metrics, including read length distribution, quality scores, and throughput analysis. Long-read nanopore sequencing was performed to a median tumor coverage of 89.9× (range 41.8–113.2×) in the PROBLEM cohort and 92.5× (range 45.8–128.5×) in the compound-variant cohort, with matched normals sequenced to a median of 36.6×. Median read N50 was 9.5 kb for PROBLEM tumors and 9.1 kb for compound-variant tumors, providing adequate read lengths for structural variant detection and haplotype phasing, and sufficient depth for calling single-nucleotide variants (SNVs), insertions/deletions (indels), and structural variants (SVs), including subclonal variants.

Read alignment to GRCh38 was performed using *minimap2* (v2.28, RRID:SCR_018550)^48^ with long-read specific parameters (-ax map-ont). Variant calling employed *Clair3* (RRID:SCR_026063)^49^ for germline variants and *ClairS* (RRID:SCR_026103) for somatic variants, both optimized for R10.4.1 chemistry (model r1041_e82_400bps_hac_v420). Variant filtering and normalization were performed with *bcftools* (v1.20, RRID:SCR_005227).

Haplotype phasing was conducted using *WhatsHap* (v2.7, RRID:SCR_025319) on matched normal samples using high-quality SNVs only, then this phasing information was applied to tumor BAM files. Read-based haplotype phasing achieved a median phase-block N50 of 92 kb in PROBLEM tumors and 350 kb in compound-variant tumors; matched normals, which retain genome-wide heterozygosity, phased to substantially longer blocks (median 771 kb). Of the compound-variant cases, 15 were sequenced by whole genome and 17 by adaptive (targeted) sampling; phase-block N50 is reported only for whole-genome samples, as adaptive sampling does not yield genome-wide phasing. All per-sample long-read statistics are provided in **Table S1**.

Structural variant detection was performed with *Severus* (v1.5)^50^, and initial calls were filtered against a curated database of recurrent technical artifacts observed in our cohort (by collapsing breakpoints to a window of 100bp). Copy number analyses on long-read samples were adapted from the methodology of *cnatools*^38–40^. Briefly, zygosity data calculated from single nucleotide polymorphism (SNP) variant allelic fraction (VAF) and log-coverage-ratio (tumor/normal) was segmented using circular binary segmentation algorithm. Ploidy and purity were jointly estimated by expectation-maximization (EM) algorithm. Methylation analysis was enabled during basecalling for 5mC and 5hmC detection (5mCG_5hmC model), with downstream analysis using *modkit* (v0.4.4, RRID:SCR_028163) for differential methylation calling and allele-specific methylation analysis. Variant annotation was carried out with *snpEff* (RRID:SCR_005191)^51^, *VEP* (RRID:SCR_007931)^52^, and *SpliceAI (*RRID:SCR_026278).^12^ The entire pipeline was automated and implemented on the Google Cloud Platform, utilizing scalable computing resources to ensure completion within clinical timelines (**Figure S1, Figure 1C**).

### Initial Mutational Signature Analyses to Nominate Patients With Genomic Instability From Short-Read Sequencing

Since patients from the PROBLEM cohort were sequenced using whole-exome sequencing and targeted panels^38–40^, we focused on mutational signatures applicable to these platforms rather than those requiring whole-genome sequencing (WGS). Single nucleotide signature analysis was performed using *SigProfilerAssignment* (v0.2.2, RRID:SCR_026899)^53^ with COSMIC (v2, RRID:SCR_002260) reference signatures in 96 trinucleotide contexts. Homologous recombination deficiency (HRD) assessment was further supplemented by R package *scarHRD*^54^. This approach uses short-read-derived copy number and zygosity data to calculate composite scores from HRD-LOH (loss of heterozygosity), large-scale transitions (LST), and telomeric allelic imbalances (TAI).^55^ In addition to COSMIC signatures, mismatch repair deficiency (MMRD) status was assessed using *MSISensor* (v0.6, RRID:SCR_006418).^56^

Overall, we used the following criteria to nominate if a patient possessed HRD, MMRD, or focal tandem duplication (FTD) phenotypes:

- MMRD inclusion criteria required either: (1) COSMIC v2 signatures 6, 15, 20, or 26 (if any of these signatures contributed ≥20%)^57^ with tumor mutation burden >10 mutations/Mb, or (2) MSISensor score >10%.
- HRD classification required: (1) scarHRD composite score ≥60 (slightly higher than the standard ≥42 threshold to account for the lower resolution of targeted sequencing^58^), or (2) combined contribution of COSMIC v2 signatures 3 and 8 ^13^ ≥30% with tumor mutation burden ≥2.5 mutations/Mb.
- FTD phenotype was defined by ≥20 focal tandem duplications (0.3-3Mb) inferred from short-read copy number data.^59^

### Secondary Mutational Signature Analyses Post-Nanopore Sequencing

Since nanopore sequencing completely resolved MMRD and FTD cohorts (**Figure 3B**), secondary mutational signature analyses were mainly focused on HRD. Unlike WES and targeted panels where signature analyses are restricted to mutational and copy number signatures, post-nanopore sequencing enables utilization of indels and structural variants. Specifically, we screened for the presence of ID6 signature (deletions with microhomology), ID8 signature (deletions at repetitive sequences), structural variant signature SV3 (short tandem duplications between 1-100Kb), and SV5 (short non-clustered deletions between 1-100Kb), which have been associated with HRD. ^15,60^ Indel and structural variant signature assignments were performed with *SigProfilerExtractor* (RRID:SCR_023121)^61^ and *SigProfilerAssignment*.^53^ We also applied the well-established HRDetect algorithm^13^ implemented in the R package *signature.tools.lib*.^14^

Based on literature reviews and clinical practices, credible HRD cases required: (1) HRDetect score >0.7 with presence of ID6 signature or SV3/SV5 signatures^15,60^, (2) HRDetect scores 0.5- 0.7 with (cryptic) double hits identified by long-read sequencing, or (3) exclusion of cases with isolated ID8 signatures and HRDetect scores < 0.7 as likely representing alternative DNA repair deficiencies or other etiologies.^60,62^ Overall, this analysis identified that only 3 of 14 unresolved cases were credibly HRD (**Figure 3B**).

### Germline-somatic co-occurrence analysis

To assess the extent to which somatic variants co-occur in *cis* with protein-altering germline variants, we performed a systematic germline-somatic co-occurrence analysis across 120 curated hereditary cancer predisposition genes in the matched tumor-normal cohort. Germline variants were called from matched normal samples and restricted to protein-altering variants, then stratified into three evidence tiers: confirmed pathogenic (ClinVar [RRID:SCR_006169] “Pathogenic” or “Likely pathogenic”), high-confidence variants of uncertain significance (CADD ≥ 25 and absent from population databases), and unannotated damaging variants (CADD ≥ 20 and absent from population databases). Population frequency was assessed using gnomAD [RRID:SCR_014964], and variants present in population databases above a rare-allele threshold were excluded. Qualifying germline variants were matched against somatic SNVs called in the corresponding tumor within the same gene to identify germline-somatic pairs. For each pair, the inter-variant genomic distance was computed, and the fraction of pairs exceeding the 500 bp short-read phasing limit was determined. This analysis identified 41 germline-somatic pairs across 39 patients and 27 genes.

### Data Visualization

Genomic data inspection and visualization were performed using IGV (v2.19.1, RRID:SCR_011793)^63^ and JBrowse^64^ (v3.6.3, RRID:SCR_001004) for genome browsing and alignment visualization. Methylation analysis utilized Methylartist (v1.5.0, RRID:SCR_028164)^65^ for allele-specific methylation plotting. Structural variant visualization employed Ribbon (v2.0)^66^ for chromosomal rearrangement plots. Statistical analysis and data visualization used R packages including ggplot2 (v3.5.2, RRID:SCR_014601)^67^, ComplexHeatmap^68^ (v2.13.1, RRID:SCR_017270), and pheatmap (RRID:SCR_016418, v1.0.12). Scientific illustrations were created using BioRender (biorender.com, RRID:SCR_018361).

## Data and Code Availability

Deidentified nanopore sequencing data supporting the conclusions of this study are available in the database of Genotypes and Phenotypes (dbGaP) under accession number phs000673. RNA- sequencing data from the NOTCH1 functional assays have been deposited in the NCBI Sequence Read Archive under BioProject accession PRJNA1484643 and will be made publicly available upon publication. Additional supporting data are available from the corresponding authors upon reasonable request.

## Data Availability

Deidentified nanopore sequencing data supporting the conclusions of this study are available in the database of Genotypes and Phenotypes (dbGaP) under accession number phs000673. Novel pathogenic germline variants identified in this study are being submitted to ClinVar. Additional supporting data are available from the corresponding authors upon reasonable request.

## Acknowledgements

This work was funded by the National Cancer Institute (NCI) Prostate SPORE Grant (P50- CA186786, to A.M. Chinnaiyan), NCI Early Detection Research Network (U2C-CA271854, to A.M. Chinnaiyan), NCI Research Specialist Award (NCI 50CA293826, to Y-M. Wu), and NCI Outstanding Investigator Award (R35-CA231996, to A.M. Chinnaiyan). A.M. Chinnaiyan is a Howard Hughes Medical Institute Investigator, Alfred Taubman Scholar, and American Cancer Society Professor.

We extend our gratitude to Stephanie Miner for editorial assistance, to Jin Chen for IT logistics, and to Kayla Muschong for support in coordinating the query of clinical information. The authors used Claude (Anthropic) to revise author-written text for clarity, grammar, and readability. No text, figures, or other content in this manuscript was generated by artificial intelligence. All authors reviewed and edited the revised text, confirmed that its meaning was unaltered, and take full responsibility for the content of this publication. Finally, we thank the patients and their families for their participation in this study and their invaluable contributions to the advancement of medical knowledge.

## Abbreviations

FTD: focal tandem duplication
HRD: homologous recombination deficiency
LOH: loss of heterozygosity
MMRD: mismatch repair deficiency
PARPi: poly(ADP-ribose) polymerase inhibitor
SNP: single-nucleotide polymorphism
SNV: single-nucleotide variant
SV: structural variant
WES: whole-exome sequencing.

