## Supplementary Tables and Figures for "Long-Read Haplotype Phasing Resolves Allelic Configuration as a Missing Layer of Precision Oncology": Supplementary Figures.pdf

Figure S1

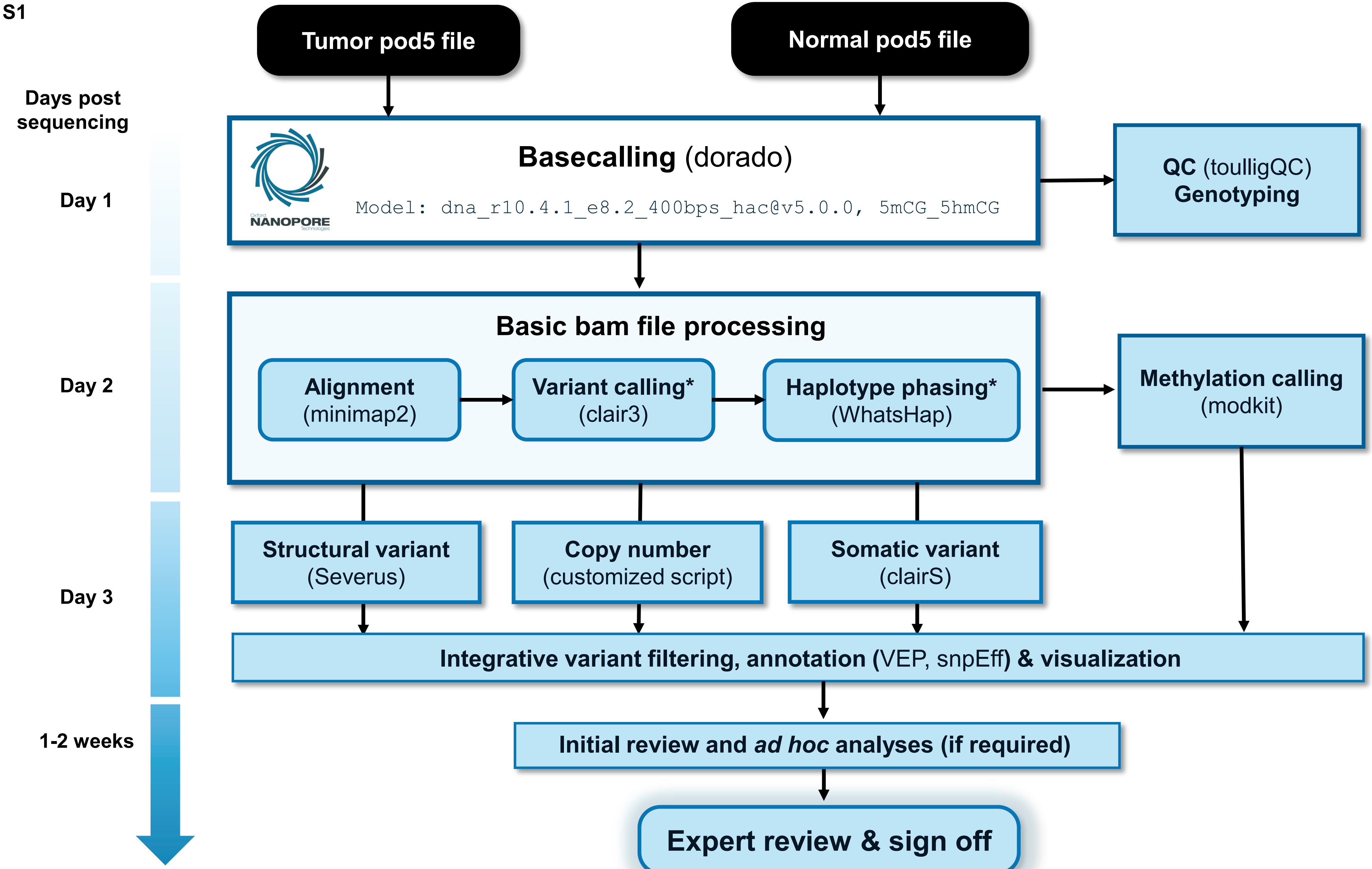

Tumor pod5 file
Normal pod5 file
Days post sequencing
Day 1
Day 2
Day 3
1-2 weeks
Basecalling (dorado)
Model: dna_r10.4.1_e8.2_400bps_hac@v5.0.0, 5mCG_5hmCG
NANOPORE
QC (toulligQC)
Genotyping
Basic bam file processing
Alignment (minimap2)
Variant calling* (clair3)
Haplotype phasing* (WhatsHap)
Methylation calling (modkit)
Structural variant (Severus)
Copy number (customized script)
Somatic variant (clairS)
Integrative variant filtering, annotation (VEP, snpEff) & visualization
Initial review and ad hoc analyses (if required)
Expert review & sign off

**Figure S1. Long-read bioinformatics pipeline.**

Comprehensive bioinformatics workflow for clinical long-read sequencing analysis. The pipeline encompasses high-accuracy model-based basecalling using Dorado (Oxford Nanopore), quality control assessment, read alignment to GRCh38 using minimap2, variant calling with Clair3 and ClairS, structural variant detection using Severus, haplotype phasing with WhatsHap, and methylation analysis with modkit for 5mC and 5hmC detection (**Methods**). Tasks marked with asterisks are performed only on normal samples, unless the analysis is tumor-only.

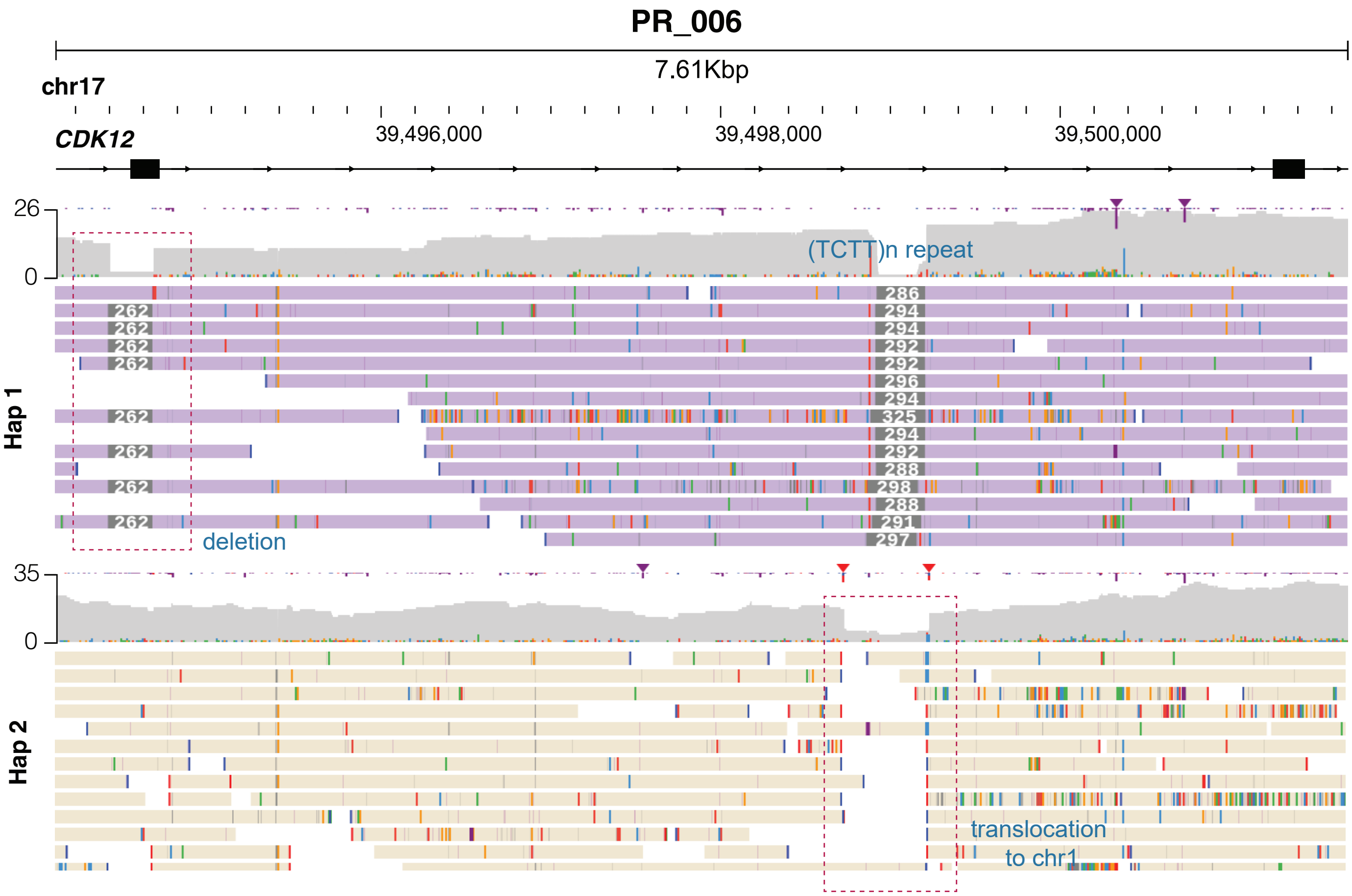


**Figure S2. An additional example of cryptic FTD mechanism.**

JBrowse visualization showing biallelic *CDK12* inactivation in case PR_006. One allele contains a 262bp deletion partially disrupting exon 5; haplotype phasing revealed a reciprocal copy-neutral translocation to chromosome 1 on the opposing allele.

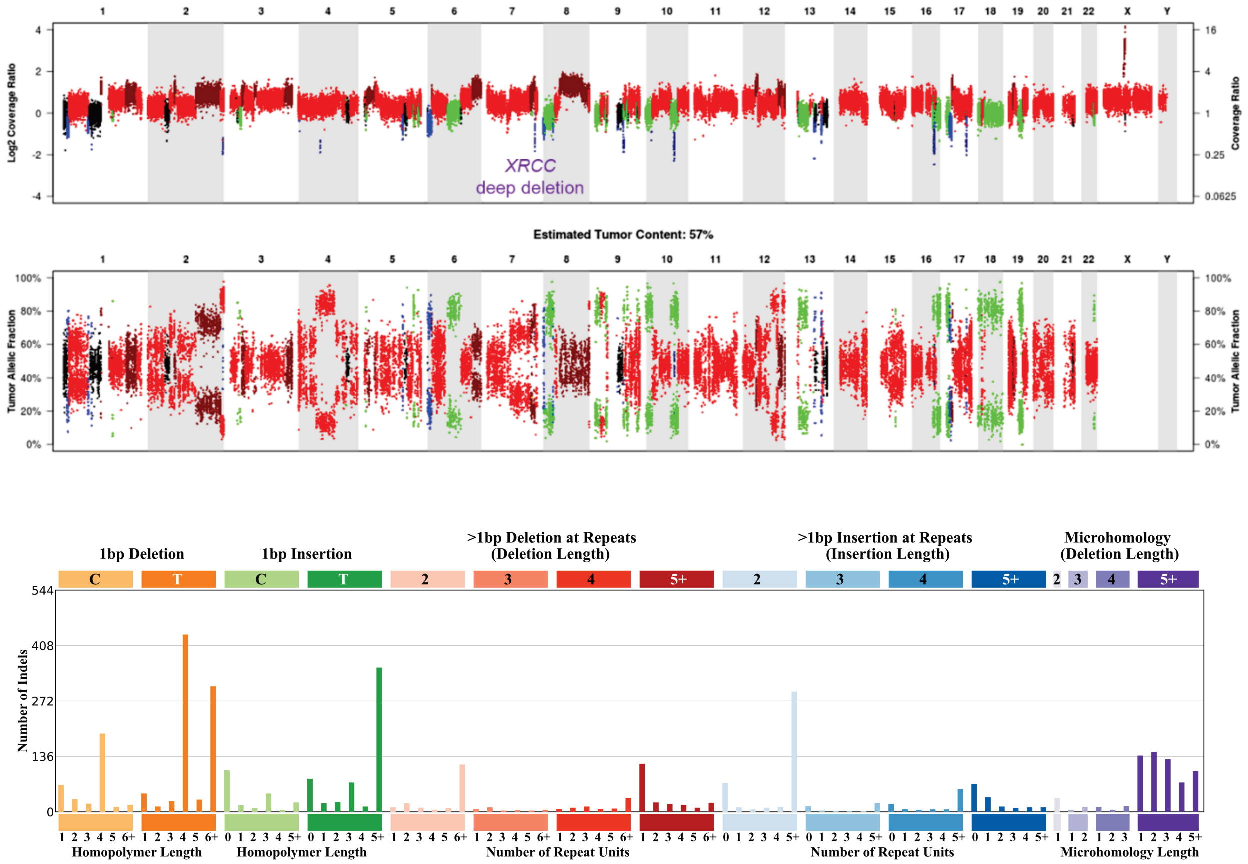


**Figure S3. Secondary analysis resolves cryptic homologous recombination deficiency through *XRCC2* deep deletion.**

A patient with prostate cancer and an elevated scarHRD score lacked an identifiable causative alteration, leaving the molecular basis of HRD unresolved in the initial clinical report. Long-read nanopore sequencing resolved this cryptic case by independently corroborating HRD-associated mutational signatures and identifying a deep deletion encompassing *XRCC2*, a core homologous recombination repair gene, as the likely driver of genomic instability. **Top:** Copy-number analysis demonstrates deep deletion of *XRCC2*. **Bottom:** Indel signatures derived from nanopore whole-genome sequencing demonstrate enrichment of deletions with microhomology (purple), a characteristic feature of HRD.

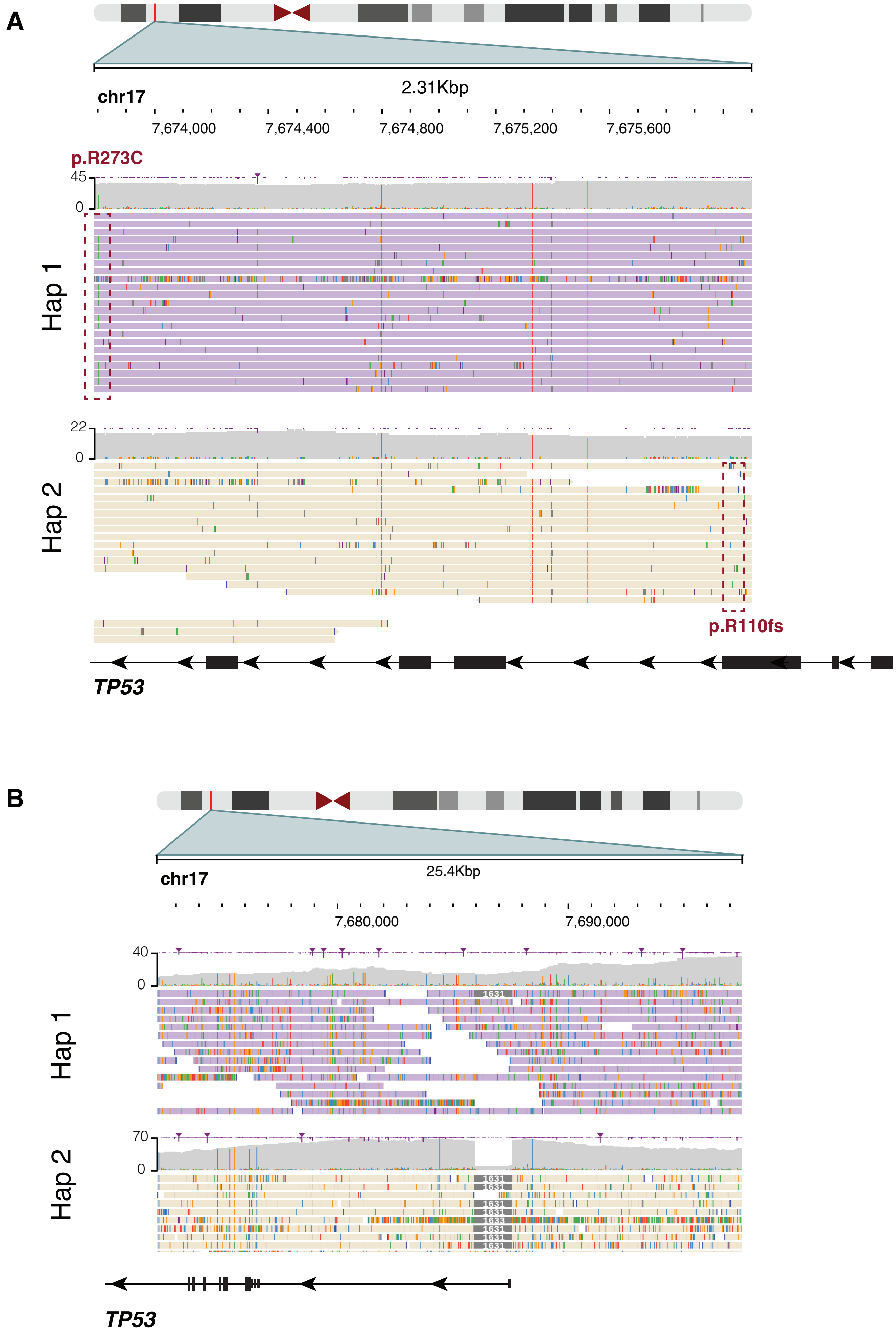

A
2.31Kbp
chr17
7,674,000
7,674,400
7,674,800
7,675,200
7,675,600
p.R273C
45
0
Hap 1
22
0
Hap 2
p.R110fs
TP53
B
chr17
25.4Kbp
7,680,000
7,690,000
40
0
Hap 1
70
0
Hap 2
TP53

**Figure S4. Long-read sequencing resolves biallelic *TP53* inactivation beyond the instability-signature gene set.**

**(A)** A metastatic castration-resistant prostate cancer harboring two pathogenic *TP53* variants (p.R273C and p.R110fs) located ~2.3 kb apart. Read-level haplotype phasing placed the variants on opposite alleles, establishing biallelic inactivation *in trans*. Because the two variants lie ~2.3 kb apart, beyond the span of short reads or read-pairs, their phase could not be determined by short-read sequencing.

**(B)** A breast cancer in which long-read sequencing identified a focal deletion affecting the first exon of *TP53* on a background of chromosome 17p loss of heterozygosity, together producing biallelic *TP53* inactivation.

### Figure S5

#### Familial Breast and Ovarian Cancer

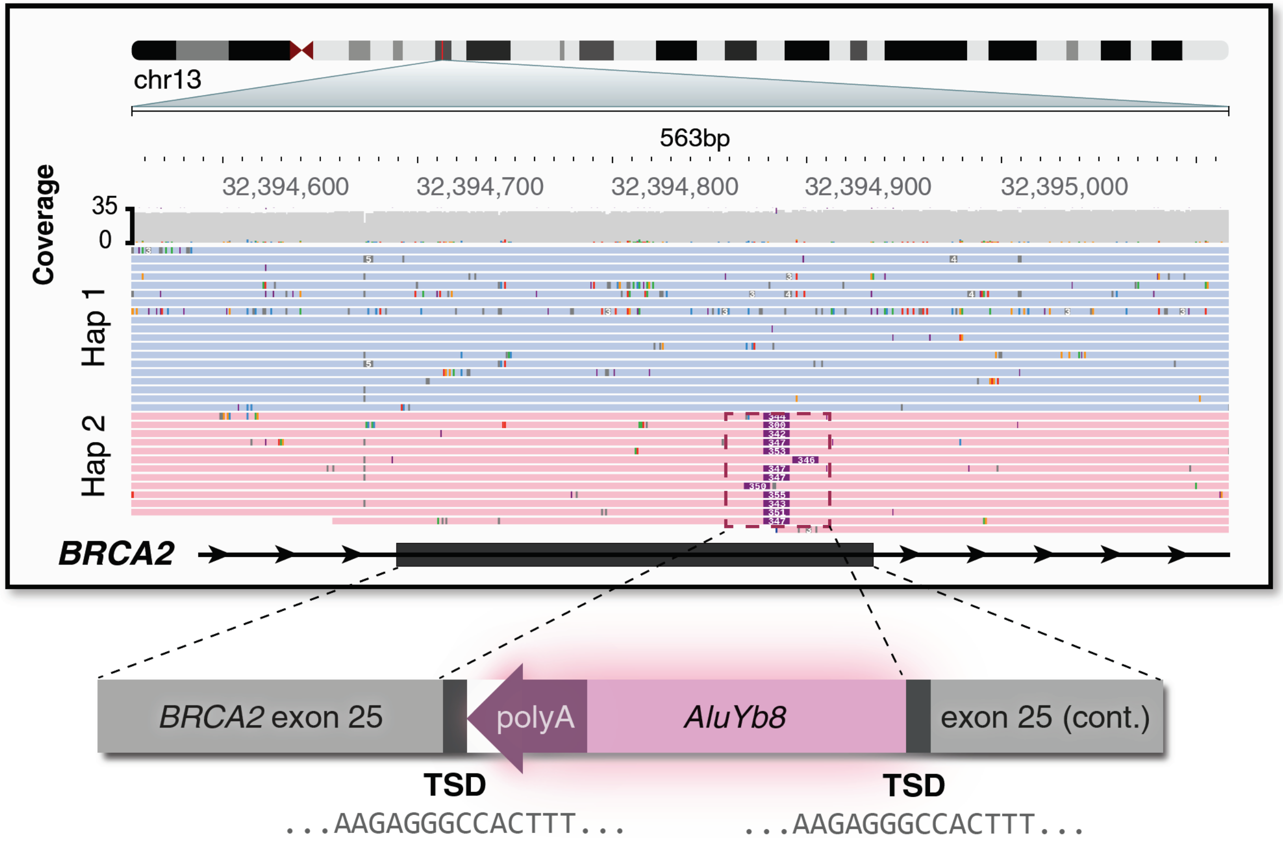


**Genomic context of ALU insertion**

```
CCTTCTTACTTTATTTGCTGGAGATTTTTCTGTGTTTTCTGCTAGTCCAAAAGAGGGCCA
CTTTCTTTTTTTTTTTTTTTTTTTTTTTTTTTTTTTTTTTTTTTTTTTTTTTTTGAGACGG
AGTCTCGCTCTGTCACCCAGGCCGGACTGCGGACTGCAGTGGCGCAATCTCGGCTCACTG
CAAGCTCCGCTTCCCGGGTTCACGCCATTCTCCTGCCTCAGCCTCCCGAGTAGCTGGGAC
TACAGGCGCCCGCCACCGCGCCCGGCTAATTTTTGTATTTTTAGTAGAGACGGGGTTTC
ACCTTGTTAGCCAGGATGGTCTCGATCTCCTGACCTCATGATCCACCCGCCTCGGCCTCC
CAAAGTGCTGGGATTACAGGCGTGAGCCACCGCGCCCGGCCAAGAGGGCCACTTTCAAGA
GACATTCAACAAAATGAAAAATACTGTTGAGGTAAGGTTACTTTTCAGCATCACCACACA
```

- Target site duplication
- ALU
- Poly(A) tail
- *BRCA2* native sequence

***AluYb8* subfamily | 318 bp | 99% identity**

| Score | Expect | Identities | Gaps | Strand |
|---|---|---|---|---|
| 582 bits(315) | 4e-171 | 317/318(99%) | 0/318(0%) | Plus/Plus |

```
Query  1    GGCCGGGCGCGGTGGCTCACGCCTGTAATCCCAGCACTTTGGGAGGCCGAGGCGGGTGGA  60
            ||||||||||||||||||||||||||||||||||||||||||||||||||||||||||||
Sbjct  1    GGCCGGGCGCGGTGGCTCACGCCTGTAATCCCAGCACTTTGGGAGGCCGAGGCGGGTGGA  60

Query  61   TCATGAGGTCAGGAGATCGAGACCATCCTGGCTAACAAGGTGAAACCCCGTCTCTACTAA  120
            ||||||||||||||||||||||||||||||||||||||||||||||||||||||||||||
Sbjct  61   TCATGAGGTCAGGAGATCGAGACCATCCTGGCTAACAAGGTGAAACCCCGTCTCTACTAA  120

Query  121  AAATACAAAAAATTAGCCGGGCGCGGTGGCGGGCGCCTGTAGTCCCAGCTACTCGGGAGG  180
            ||||||||||||||||||||||||||||||||||||||||||||||||||||||||||||
Sbjct  121  AAATACAAAAAATTAGCCGGGCGCGGTGGCGGGCGCCTGTAGTCCCAGCTACTCGGGAGG  180

Query  181  CTGAGGCAGGAGAATGGCGTGAACCCGGGAAGCGGAGCTTGCAGTGAGCCGAGATTGCGC  240
            ||||||||||||||||||||||||||||||||||||||||||||||||||||||||||||
Sbjct  181  CTGAGGCAGGAGAATGGCGTGAACCCGGGAAGCGGAGCTTGCAGTGAGCCGAGATTGCGC  240

Query  241  CACTGCAGTCCGCAGTCCGGCCTGGGTGACAGAGCGAGACTCCGTCTCaaaaaaaaaaaa  300
            |||||||||||||||||||||||||| |||||||||||||||||||||||||||||||||
Sbjct  241  CACTGCAGTCCGCAGTCCGGCCTGGGCGACAGAGCGAGACTCCGTCTCAAAAAAAAAAAA  300

Query  301  aaaaaaaaaaaaaaaaaa  318
            ||||||||||||||||||
Sbjct  301  AAAAAAAAAAAAAAAAAA  318
```

**Figure S5. *AluYb8* retrotransposon insertion in *BRCA2*.**

Genomic sequence analysis details the structural features of a pathogenic *AluYb8* insertion within *BRCA2* exon 25, detected in a woman in her 60s with a strong family history of breast and ovarian cancer and prior negative commercial multi-gene panel testing. Target site duplications and insertion boundaries are shown. BLAST alignment results demonstrate 99.7% sequence identity to the *AluYb8* consensus sequence, differing by only a single nucleotide, consistent with a relatively recent insertion event. Long-read sequencing enabled unambiguous detection of this insertion, which is invisible to standard short-read sequencing.

Figure S6

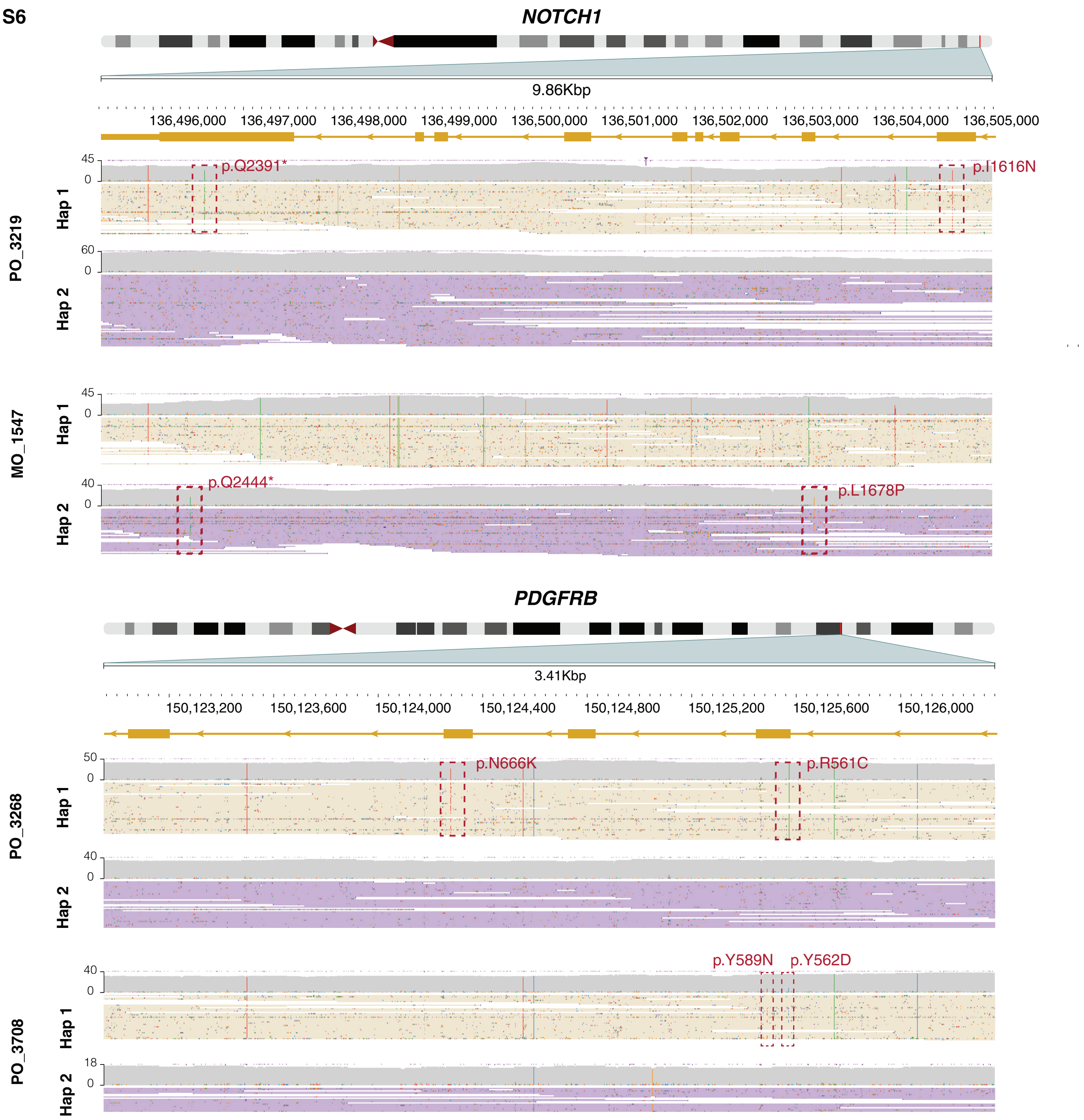

NOTCH1
9.86Kbp
136,496,000
136,497,000
136,498,000
136,499,000
136,500,000
136,501,000
136,502,000
136,503,000
136,504,000
136,505,000
45
0
p.Q2391*
p.I1616N
60
0
PO_3219
Hap 1
Hap 2
45
0
40
0
p.Q2444*
p.L1678P
MO_1547
Hap 1
Hap 2
PDGFRB
3.41Kbp
150,123,200
150,123,600
150,124,000
150,124,400
150,124,800
150,125,200
150,125,600
150,126,000
50
0
p.N666K
p.R561C
40
0
PO_3268
Hap 1
Hap 2
40
0
p.Y589N
p.Y562D
18
0
PO_3708
Hap 1
Hap 2

**Figure S6. JBrowse screenshots confirming *cis* configuration of compound *NOTCH1* and *PDGFRB* variants.**

JBrowse visualization demonstrates definitive *cis* configuration of compound variants in *NOTCH1* (affecting heterodimerization and PEST domains) and *PDGFRB* (affecting the juxtamembrane regulatory regions). Long-read sequencing enables unambiguous determination of variant phasing across kilobase-scale distances that are irresolvable by short-read methods. Individual long reads spanning both variant positions are shown for representative cases from each gene.

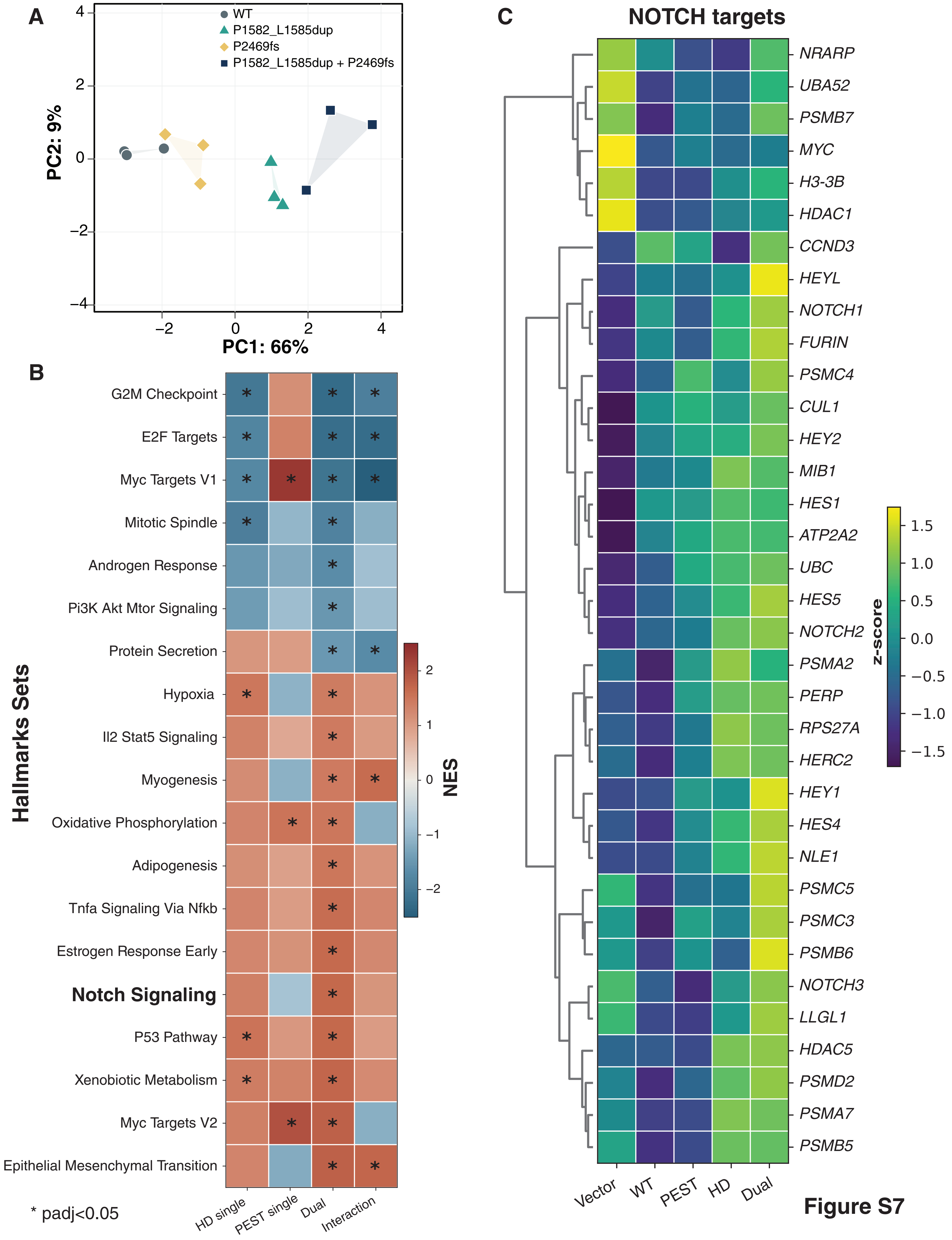
A
WT
P1582_L1585dup
P2469fs
P1582_L1585dup + P2469fs
PC2: 9%
PC1: 66%
B
G2M Checkpoint
E2F Targets
Myc Targets V1
Mitotic Spindle
Androgen Response
Pi3K Akt Mtor Signaling
Protein Secretion
Hypoxia
Il2 Stat5 Signaling
Myogenesis
Oxidative Phosphorylation
Adipogenesis
Tnfa Signaling Via Nfkb
Estrogen Response Early
Notch Signaling
P53 Pathway
Xenobiotic Metabolism
Myc Targets V2
Epithelial Mesenchymal Transition
Hallmarks Sets
NES
HD single
PEST single
Dual
Interaction
* padj<0.05
C
NOTCH targets
NRARP
UBA52
PSMB7
MYC
H3-3B
HDAC1
CCND3
HEYL
NOTCH1
FURIN
PSMC4
CUL1
HEY2
MIB1
HES1
ATP2A2
UBC
HES5
NOTCH2
PSMA2
PERP
RPS27A
HERC2
HEY1
HES4
NLE1
PSMC5
PSMC3
PSMB6
NOTCH3
LLGL1
HDAC5
PSMD2
PSMA7
PSMB5
z-score
Vector
WT
PEST
HD
Dual

**Figure S7**

**Figure S7. Compound *cis NOTCH1* mutations elicit an intensified transcriptional program relative to single-site mutants.**

**(A)** Principal component analysis of RNA-seq profiles from HEK293 cells expressing wild-type NOTCH1 (WT) or the indicated ACC-associated mutants: the HD-domain mutant P1582_L1585dup, the PEST-domain mutant P2469fs, or the compound *cis* allele (P1582_L1585dup + P2469fs). PCA was computed on the top 500 most variable genes (*NOTCH1* excluded) after variance-stabilizing transformation; empty-vector controls were omitted. Each point is a biological replicate (n = 3 per genotype); symbols/colors denote genotype and shaded regions show the convex hull of each group's replicates. Samples separate primarily along PC1 (66% of variance) in the order WT → single mutants → compound mutant, with the compound allele displaced furthest from WT.

**(B)** Gene set enrichment analysis (GSEA, fgsea) of Hallmark gene sets across four contrasts: each single mutant versus WT (HD single, PEST single), the compound mutant versus WT (Dual), and the genotype interaction term (Interaction). Shown are all Hallmark sets significantly enriched in the Dual contrast (padj < 0.05). Color indicates normalized enrichment score (NES; red, up; blue, down relative to WT); asterisks mark padj < 0.05. The Interaction column tests for non-additivity (the deviation of the compound mutant from the sum of the two single-mutant effects, modeled as the HD:PEST term in a 2×2 factorial design); enrichment in this column indicates a synergistic (non-additive) program. Genes were ranked by the Wald statistic for each contrast.

**(C)** Hierarchically clustered heatmap of NOTCH pathway genes across genotypes, including canonical targets and leading-edge genes from the significant NOTCH GSEA sets. Values are per-genotype mean expression z-scores (row-scaled across the five groups: Vector, WT, PEST, HD, Dual). Rows were clustered by Euclidean distance (Ward linkage); columns are shown in fixed order. Canonical NOTCH targets (e.g. *HES4*, *HES5*, *HEY1*, *HEY2*, *HEYL*, *CCND3*) are induced progressively from WT to the compound mutant.
