## Supplementary Tables and Figures for "Long-Read Haplotype Phasing Resolves Allelic Configuration as a Missing Layer of Precision Oncology": Table S2.pdf

**Table S2. Resolved Mismatch Repair Deficiency (MMRD) Cases with Cryptic Double Hits.**

| Patient | Cancer Type | First Hit | Second Hit |
|---|---|---|---|
| PR_031 | Prostate cancer (mCRPC) | *MSH2* deletion | *MSH2* inversion |
| PR_030 | Prostate cancer (mCRPC) | *MSH2* deletion | *MSH2* translocation |
| PR_032 | Neuroendocrine prostate cancer | *PMS2* deletion | *PMS2* complex rearrangement |
| PR_028 | Breast cancer | *MLH1* promoter methylation | *MLH1* p.Gly244Val (rs63750303)§ |
| PR_029 | Breast cancer | *MLH1* promoter methylation | *MLH1* promoter methylation |

§ Variant identified by short-read sequencing
