## Supplementary Tables and Figures for "Long-Read Haplotype Phasing Resolves Allelic Configuration as a Missing Layer of Precision Oncology": Table S3.pdf

**Table S3. Resolved Focal Tandem Duplication (FTD) Cases with Cryptic Double Hits.**

| Patient | Cancer Type | First Hit | Second Hit |
|---|---|---|---|
| PR_002 | Prostate cancer (mCRPC) | *CDK12* tandem duplication | *CDK12* translocation |
| PR_003 | Prostate cancer (mCRPC) | *CDK12* focal deep deletion | LOH |
| PR_004 | Prostate cancer (mCRPC) | *CDK12* complex rearrangement | *CDK12* 3'UTR deletion |
| PR_005 | Prostate cancer (mCRPC) | *CDK12* inversion | LOH |
| PR_006 | Prostate cancer (mCRPC) | *CDK12* exon 5 partial deletion | *CDK12* translocation |
| PR_007 | Prostate cancer (mCRPC) | *CDK12* translocation | *CDK12* translocation |
| PR_001 | Serous ovarian cancer | *CDK12* alternative exon usage | LOH |
