## Supplementary Tables and Figures for "Long-Read Haplotype Phasing Resolves Allelic Configuration as a Missing Layer of Precision Oncology": Table S4.pdf

**Table S4. Resolved Homologous Recombination Deficiency (HRD) Cases with Cryptic Drivers.**

| Patient | Cancer | First Hit | Second Hit |
|---|---|---|---|
| PR_008 | Breast cancer | *BRCA1* methylation | *BRCA1* methylation |
| PR_012 | Breast cancer | *BRCA1* methylation | *BRCA1* methylation |
| PR_013 | Breast cancer | *BRCA1* methylation | *BRCA1* methylation |
| PR_014 | Breast cancer | *BRCA1* methylation | *BRCA1* methylation |
| PR_017 | Breast cancer | *BRCA1* methylation | *BRCA1* methylation |
| PR_018 | Breast cancer | *BRCA1* methylation | *BRCA1* methylation |
| PR_020 | Breast cancer | *BRCA1* methylation | *BRCA1* methylation |
| PR_023 | Breast cancer | *BRCA1* methylation | *BRCA1* methylation |
| PR_027 | Breast cancer | *BRCA1* methylation | *BRCA1* methylation |
| PR_016 | Breast cancer | *BRCA1* methylation | *BRCA1* methylation |
| PR_022 | Breast cancer | *BRCA1* internal tandem dup | LOH |
| PR_019 | Breast cancer | *BARD1* partial deletion | - |
| PR_021 | Breast cancer | *BARD1* partial deletion† | *BARD1* (NM_000465.4:c.159-5A>G) |
| PR_009 | Breast cancer | *PALB2* p.Gln39Ter§ | *PALB2* (NM_024675.4:c.2835-12T>G)† |
| PR_010 | Breast cancer | *RAD51C* methylation | LOH |
| PR_011 | Ovarian cancer | *BRIP1* internal deletion† | LOH |
| PR_015 | Prostate cancer | *BRCA2* inversion | *BRCA2* deletion |
| PR_024 | Prostate cancer | *BRCA2* inversion | *BRCA2* p.Thr2968fs †§ |
| PR_025 | Prostate cancer | *BRCA2* internal deletion | *LOH* |
| PR_026 | Prostate cancer | *BRCA2* complex SV | *LOH* |

†Germline variants

§Variants identified by short-read sequencing
